# Prevalence of cognitive impairment following chemotherapy treatment for breast cancer: A systematic review and meta-analysis

**DOI:** 10.1101/2021.08.17.21262190

**Authors:** Alexandra L Whittaker, Rebecca P George, Lucy O’Malley

**Author notes:** MeSH key words: Chemotherapy-Related Cognitive Impairment; Psycho-Oncology; Breast Neoplasms; Chemotherapy, Adjuvant.

## Abstract

Breast cancer survival rates have markedly improved. Consequently, survivorship issues have received increased attention. One common sequela of treatment is chemotherapy-induced cognitive impairment (CICI). CICI causes a range of impairments that can have a significant negative impact on quality of life. Knowledge of the prevalence of this condition is required to inform survivorship plans, and ensure adequate resource allocation and support is available for sufferers.

**Objective:** To estimate the prevalence of cognitive impairment following chemotherapy treatment for breast cancer.

**Methods:** Medline, Scopus, CINAHL and PSYCHInfo were searched for eligible studies which included prevalence data on CICI, as ascertained though the use of self-report, or neuropsychological tests. Methodological quality of included studies was assessed. Findings were synthesised narratively, with meta-analyses being used to calculate pooled prevalence when impairment was assessed by neuropsychological tests.

**Results and discussion:** The review included 52 studies. Time-points considered ranged from the chemotherapy treatment period to greater than 10 years after treatment cessation. Summary prevalence figures (across time-points) using self-report, short cognitive screening tools and neuropsychological test batteries were 44%, 16% and 21-34% respectively (very low GRADE evidence).

**Conclusion:** Synthesised findings demonstrate that 1 in 3 breast cancer survivors may have clinically significant cognitive impairment. Prevalence is higher when self-report based on patient experience is considered. This review highlights a number of study design issues that may have contributed to the low certainty rating of the evidence. Future studies should take a more consistent approach to the criteria used to assess impairment. Larger studies are urgently needed.

**Summary of Findings Table:** 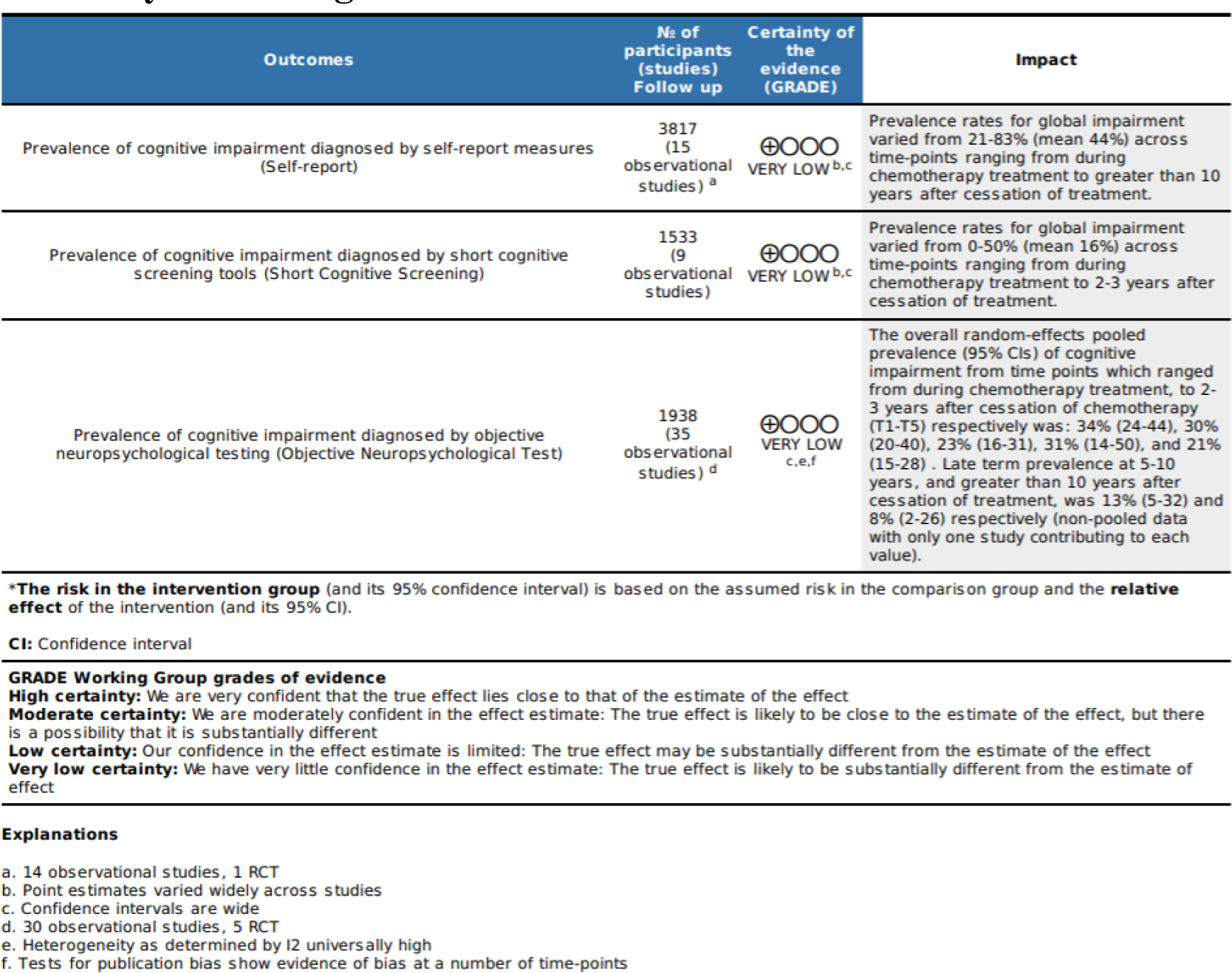

## Background

In women, breast cancer is the most frequently diagnosed cancer, with an estimated 2.3 million new cases globally per year. [1] Prognosis has significantly improved over time, with current 5-year survival rates of around 90% and 10-year survival at 80%.[1] This improvement has likely been brought about largely through the ability to characterise cancer subtypes, enabling development of targeted agents, increased use of personalised medicine and consequently improved treatment success. [1] Concurrently, improvements in surgical resection methods have also been seen. [2] As a result, the number of breast cancer patients living beyond a diagnosis has grown significantly, and survivorship issues are receiving more attention, with a particular focus on quality of life issues.

Breast cancer treatment generally uses a multi-modal treatment approach utilising combinations of surgery, chemotherapy, endocrine therapy and radiotherapy, dependant on disease stage and sub-type classification. [3] However, chemotherapy combined with surgery, especially in advanced stages of the disease, forms the mainstay of therapy.

Chemotherapy has been implicated as a significant contributor to cognitive impairment observed in cancer patients, both during and after cessation of treatment. The resultant condition has been termed chemotherapy-induced cognitive impairment (CICI), or colloquially, chemofog or chemobrain. [4] Whilst the exact mechanisms for this are unclear, proposed mechanisms include direct neurotoxic injury, a reduction in neurogenesis (new neuron formation), central nervous system white matter abnormalities, and neuroinflammation. [5] Most of the evidence for these theories comes from the pre- clinical literature, [5–7] or neuro-imaging studies [8, 9].

The most common neuropsychological effects that arise in CICI are impairments to visual processing, visual motor function, executive function and attention. [10] Sufferers typically report side effects with a wide range of severity, from subtle to more severe impairment, which may persist for up to 20 years post-chemotherapy treatment. [11] Impairments typically manifest as survivors feeling ‘less-sharp’, being unable to recall words, having episodes of metal confusion, and requiring more mental effort to perform everyday activities. [12] [10] As a result, CICI has the potential to significantly impact patient quality of life. Furthermore, it has been suggested that a lack of prior information, coupled with minimal validation and understanding from family and health care providers leads to patients feeling disempowered, and subsequently not receiving the emotional and rehabilitative support they may need. [13] The reported prevalence of CICI following treatment for breast cancer is somewhat variable. A commonly cited range from the narrative review of Janelsins et al. 2014 suggests that 12 – 82% of breast cancer patients will experience impairment as a result of their treatment regime. [14] This variability may be attributable to a range of patient factors such as age, [15] IQ, [16] menopausal status, [16] and education level. [17] Alternately, study design factors such as the employment of cross-sectional, versus longitudinal study designs, with the latter able to account for baseline pre-treatment cognitive function, likely influences rates. [18] Furthermore, in spite of the name, it has been shown that cognitive impairment is often present before chemotherapy treatment has commenced, potentially arising as a result of cancer itself. [19, 20] This may occur via direct effects of the tumour itself, as a result of associated co-morbidities, or due to psychological factors such as worry and fatigue. [21] Time since treatment, also likely contributes to the variance in prevalence reported. Neuroimaging has demonstrated structural and functional changes in various patient brain regions, which likely correlate with the corresponding changes seen in executive function and memory. However, these alterations also show partial recovery over time, [22, 23] presumably leading to an improvement in symptoms. Finally, prevalence rates may be influenced by the method used to assess cognitive impairment, with the three common methods employed being the ‘gold-standard’ neuropsychological testing, [24] short cognitive screening tools [25, 26] and self-report. [14]

A preliminary database search for previous systematic reviews on prevalence of CICI was conducted. One recent systematic review was sourced which reports prevalence data for cognitive impairment in breast cancer patients. [3] This review is limited in scope compared to the current review since the timeline of consideration was the breast cancer treatment period, rather than longer-term follow up points. The review authors also only included longitudinal studies, which utilized objective neuropsychological tests. Whilst the reasons for these inclusions are understandable, the present review expands on this knowledge base by evaluating both self-report and objective psychological testing to compare prevalence rates of CICI ascertained by these different testing modalities. In the current review all study designs were eligible, not just longitudinal designs. This broad inclusion does not allow the teasing apart of the contribution of chemotherapy, as opposed to cancer itself, on impairment. However, from the point of view of the patient suffering cognitive difficulties, or health-care providers and policy makers needing evidence-based information, this distinction is of little consequence; external validity of the findings is assured. Understanding the burden of CICI will help inform survivorship strategies, and drive funding priorities for future research and targeted support, such as rehabilitation options. Furthermore, by identifying CICI incidence over the longer term this will indicate the need for longer-term support to inform cancer survivorship care plans.

### Review Question

The aim of this review was to synthesise the evidence on prevalence rates of cognitive impairment following chemotherapy treatment in breast cancer survivors, taking into consideration factors such as age and time since cessation of treatment.

## Methods

The Joanna Briggs Institute guidelines on conducting prevalence reviews [27] and the PRISMA guidelines [28] were used to guide review performance and reporting. The objectives, inclusion criteria and methods of analysis for this review were specified in advance and documented in a protocol registered on PROSPERO; ref CRD42021228541.

### Search Strategy

The search strategy aimed to locate published studies in English. An initial search of Medline was undertaken to identify articles on the topic. Keywords used in the titles and abstracts of relevant articles, and the index terms used to describe the articles were used to develop a full search strategy for Medline via Pubmed using MeSH and free text terms. The four databases were searched in December 2020 using the developed search strategies (see supplementary material S1). Key concepts used for searching were “cancer”, “chemotherapy” and “cognitive impairment”. Hand searching of reference lists was performed to identify additional studies. Studies published from database inception were eligible for inclusion. Publications were excluded if they were conference abstracts, review articles and grey literature.

### Study Selection

Following the search, all identified citations were uploaded into EndNote X8.0.1 and duplicates removed. Potentially relevant studies were retrieved in full and their citation details imported into Covidence (Veritas Health Innovation, Melbourne, Australia). Title, abstract and full text screening for assessment against the inclusion criteria for the review (see below) was performed by one reviewer (A.W), with checking of the information performed by a second reviewer (R.P.G).

### Eligibility Criteria

#### Population

Females of any age treated with systemic chemotherapeutic agents for any form of breast cancer were considered for inclusion. Patients currently in treatment or that had ceased treatment (remission) were eligible for inclusion. Studies with patients receiving other concurrent drug therapy or with other co-morbidities were eligible for inclusion with noting (see exclusion criteria). Patients of any socioeconomic status or level of education were eligible for inclusion with noting of any differences reported. Patients that were either pre-or post-menopausal were eligible for inclusion with noting.

The following groups of patients were excluded:

- patients with previous history of traumatic brain injury, neurodegenerative conditions such as Parkinson’s or Alzheimer’s or severe depressive symptoms, since these conditions affect cognition either directly or indirectly. [29]
- patients who received palliative care (due to differences in the treatment pathways and treatment regimens).[30]
- patients with metastatic CNS cancers or who have previously received CNS targeted therapy for other conditions due to the risk of direct effect on cognition as a result of the cancer/treatment [31, 32]

#### Intervention

Studies evaluating patients administered chemotherapy for breast cancer treatment were included. This included a range of common chemotherapy regimens using either sole or combination chemotherapy across any number of treatment cycles. Studies evaluating radiation and newer targeted therapies for cancer treatment were excluded. However, patients who received hormone therapy, such as tamoxifen, for prophylactic purposes following the treatment of their cancer, or accompanying local radiation therapy were eligible for inclusion with noting. This represents a deviation from the published protocol since it was discovered on screening that the majority of the studies have included these patients within their chemotherapy –treatment groups. Given that these treatment strategies are commonly used in breast cancer patients, this inclusion is justified in enhancing study external validity.

#### Condition

Studies were included if they were investigating cognitive impairment arising as a result of interventions administered for cancer as described above. This condition may be variably described as ‘chemobrain’, ‘chemofog’, chemotherapy-induced cognitive impairment (CICI), or cancer-related cognitive impairment (CRCI). Studies where cognitive decline was indicated by any one of several commonly used modalities were eligible for inclusion. These measurement modalities included self-reported measures, imaging, or objective neuropsychological testing [33].

#### Context & Study Design

Studies from any country or geographic region were eligible for inclusion with noting of location. Patients from community or clinic settings were eligible for inclusion. All study designs were eligible for inclusion. Studies that selected participants into groups based on report of cognitive decline were excluded due to the risk of sampling bias limiting the generalisability of the results.

#### Data extraction

Data were extracted from the included studies by one reviewer (A.W), with checking performed on all studies by a second reviewer using a modified extraction template in Covidence (supplementary material S2). The template was piloted on three studies to confirm that all required information was being considered. Contact with study authors was undertaken where necessary to clarify findings or seek further information. Key data extracted included the population sampled, chemotherapy and other treatment administered, timeframe since chemotherapy treatment or remission, identified confounding factors, method of cognitive testing and criteria for classifying impairment. Prevalence (%) was extracted or calculated from the available data. Where necessary, data were extracted from figures using GetData Graph Digitiser (version 2.26, S. Federov, Moscow, Russia). Prevalence estimates reported by sub-group, such as cognitive domain affected, age, or treatment were also recorded.

#### Assessment of Methodological Quality

Articles included in the study were assessed for methodological quality by one reviewer (AW), with confirmation provided by a second reviewer, using the JBI critical appraisal checklist and guidance notes developed for prevalence reviews (supplementary material S3). [34] A study was deemed to be of high quality if it scored greater than 70%, moderate quality with scores between 50-70% and low quality when scoring below 50%, as described in Dijkshoorn et al 2021 [3]. Summary graphs were created in Review Manager (RevMan) ([Computer program], Version 5.3. Copenhagen: The Nordic Cochrane Centre, The Cochrane Collaboration, 2014).

#### Data synthesis

An initial descriptive analysis of the studies was undertaken to identify relevant subgroups where studies with similar characteristics could be combined. Synthesis was undertaken narratively where there was heterogeneity in these study characteristics.

Studies were grouped for synthesis based on time point of cognitive assessment. Basic statistical analysis was performed using Megastat Excel Add-In (version 10.3, McGraw- Hill Higher Education, New York, NY).

A meta-analysis of the prevalence rates of cognitive impairment was conducted for studies that utilised objective neuropsychological tests. Many of the included studies contained repeated observations where the unit of interest was the individual rather than observation. Consequently, these cannot be combined in a standard meta-analysis, with time-point as sub-group, since they introduce a unit-of-analysis error. [35] A number of approaches to perform meta-analyses in this situation have been proposed. [36] For this review, an All Time-points Meta-analysis (ATM) method was selected with studies at the time-points of interest being analysed separately, with a qualitative comparison being made with estimates at other time-points. Analysis was performed using the MetaXL (www.epigear.com) add-in for Microsoft Excel. A pooled prevalence figure was calculated with 95% CI. Overall estimates were calculated with random effects models employing the DerSimonian & Laird method as a between study variance estimator.

Cochran’s Q (verifying the presence of heterogeneity) and the *I*^2^ statistic (amount of heterogeneity) were calculated. A quality model [37] was run as part of the meta-analysis where the weight of each study was calculated based on the previously calculated quality score. An *I*^2^ > 70% was considered substantial and these pooled data were subject to sensitivity analyses.

In a meta-analysis of prevalence, if the estimates for a study get closer to the limits of zero or one, the weight may be overestimated in the meta-analysis. Consequently, it has been recommended to transform the prevalence to a variable not constrained to the 0-1 range with an approximately normal distribution. [38] The double arcsine transformation is usually preferred [38] and was applied here. For final presentation, the pooled transformed proportions and corresponding CIs were back transformed.

Publication bias was examined using doi plots and the Luis Furuya-Kanamori asymmetry index (LFK index) in MetaXL. The doi plot is suggested to be more sensitive than the funnel plot, particularly when small numbers of studies are involved. [39]

Potential influences on prevalence estimates were investigated using subgroup analyses and meta-regression. The criteria for performing a meta-regression were derived from Cochrane guidance. [35] Meta-regression was performed using Comprehensive Meta- Analysis (Version 3, Borenstein, M., Hedges, L., Higgins, J., & Rothstein, H. Biostat, Englewood, NJ 2013). Age, cognitive impairment criteria stringency, sample size, methodological quality and geographical location were identified a priori as potential sources of variation in the prevalence estimates.

## Results

### Search Results

A total of 3010 titles were identified from the electronic searches, with 5 identified from reference lists (Figure 1). Following removal of duplicates 2194 titles remained. After title screening, 127 records remained. A further 43 records were excluded after abstract examination. Eighty-nine full-text articles were screened. Fifty-two of these met the eligibility criteria for the review, with thirty-seven being excluded at this stage (refer to supplementary material S4).

**Figure 1:**
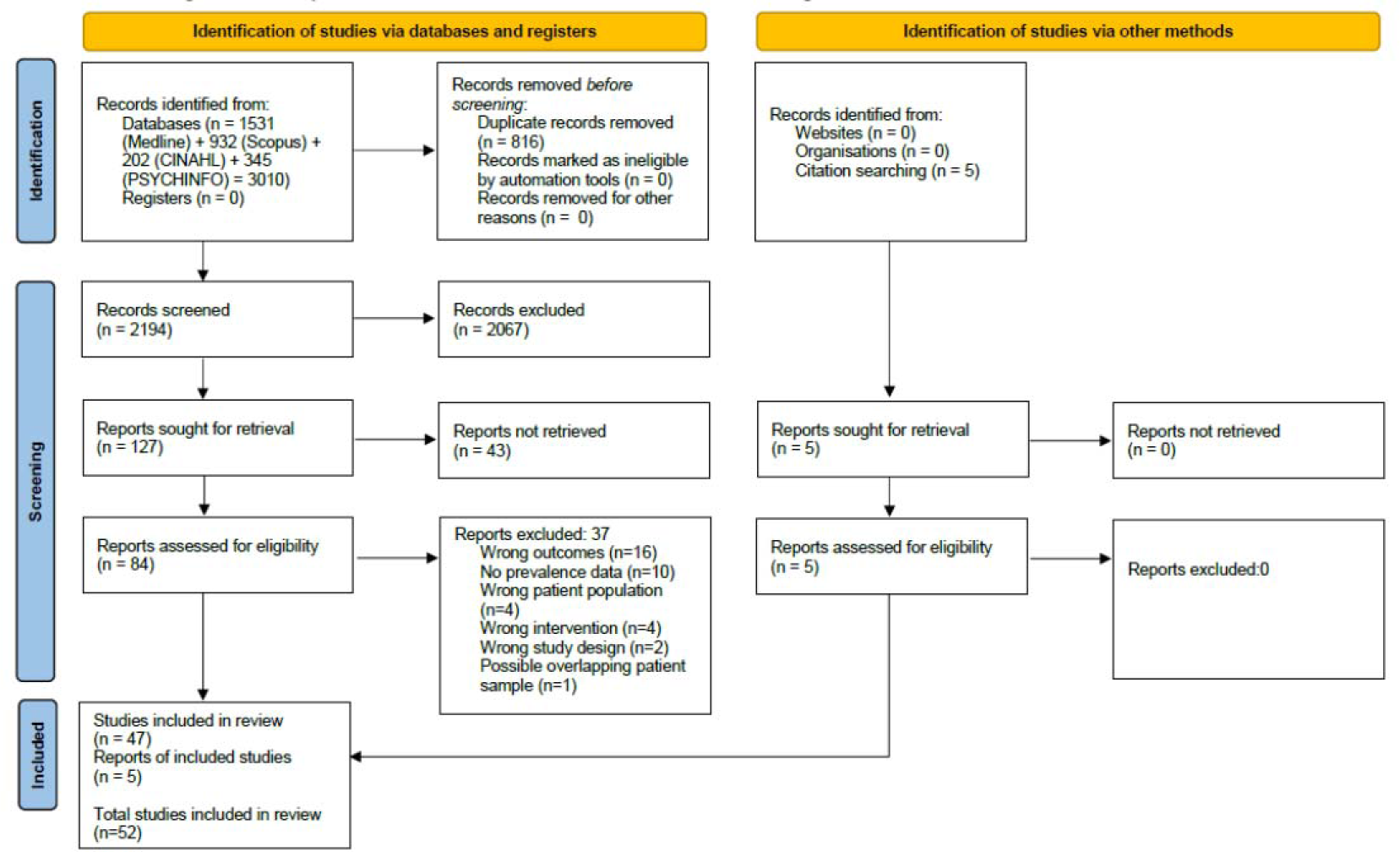
The PRISMA [28] flow diagram for the systematic review detailing the databases searched, the number of abstracts screened and the full texts retrieved

### Included Studies

The 52 included studies contributed 84 prevalence estimates across a series of time points in relation to chemotherapy treatment. It should be noted that the majority of studies did not seek to determine prevalence as a primary aim of the study, with the data being presented as one of a number of outcomes reported. Three studies did however have a focus on prevalence as expounded in their study aims. [40–42] The majority of studies adopted an observational study design (48 studies, 92%). The remainder were randomised controlled trials (RCT). The studies that employed RCT designs were evaluations where the primary study question related to differences between different chemotherapy regimes or alternate therapeutic strategies and there was no selection into groups based on presence of cognitive impairment (an exclusion criterion for this review). Out of the studies that were observational in nature, 25 (52%) were cohort designs, 20 (42%) cross-sectional and 13 (27%) prospective longitudinal. Sample sizes were generally small with a range from 20 - 1147. Studies with larger sample sizes tended to employ self-report measures rather than objective test modalities.

With the exception of two studies [43, 44], data came from countries with high-income economies (World Bank Classification). The vast majority of study data originated from the continents of North America and Europe (42% and 44% respectively). Age of study participants was generally similar across studies and to be expected given the breast cancer age risk. The pooled mean across the studies was age 52 (range of included study means: 45-71). Three studies [15, 45, 46] were specifically interested in older patients and included only women over the age of 65.

Consistent with clinical practice, women received a range of chemotherapy drugs as part of their treatment cycles across the studies. Common combinations reported were FEC (5-fluorouracil, epirubicin, and cyclophosphamide), CMF (cyclophosphamide, methotrexate and 5-fluorouracil), doxorubicin + paclitaxel, doxorubicin + cyclophosphamide. This information has not been presented in the review due to a general lack of separation of outcome data based on treatment in the primary studies, necessitating assimilation as a group. In the vast majority of studies, a significant percentage of patients, especially at later follow-up points were receiving hormonal therapy such as tamoxifen. Hormonal therapies have been shown to influence cognition [47, 48]. These agents are administered frequently and can be considered a standard part of treatment. Inclusion of these data hence supports the external validity of the review’s findings.

The included studies generally assessed cognition at similar time-points to coincide with typical phases of breast cancer treatment. Due to this similarity studies were grouped based on these time points for further assimilation, labelled T1-T7 (Refer to supplementary material S5). Data from baseline, pre-chemotherapy assessments (T0), were not extracted as part of this review since we did not have as an inclusion criterion that studies had to be longitudinal. Further, the review question relates to prevalence of impairment after breast cancer treatment, not any measured decline as a result of treatment.

The included studies utilized a range of methods for assessment of cognitive impairment. It was considered that these differences could introduce substantial heterogeneity when assimilating prevalence estimates. Hence, assimilation was undertaken based on use of three different methods; self-report measures, short cognitive screening tools, and objective neuropsychological test batteries (supplementary material S6). Fifteen studies utilized self-report measures, nine employed short cognitive screening tools and a majority of 35 used neuropsychological test batteries. Seven studies utilized two of these testing modalities [15, 49–51] [52] [53] [42]. Studies using objective cognitive tests for the most part used tests which spanned the various cognitive domains of executive function, language function, motor, processing speed, verbal and visual learning and memory, visuo-spatial function and working memory. From here on, review data is presented based on these 3 groupings of outcome assessment method (refer to Table 1, 2 and 3).

**Table 1:** Summary of included studies that used self-report measures of cognitive impairment

| Study | Study Design | Location | Sample Size | Sample Age (Range, Mean) | Assessment Method | Methodological Quality | Definition of cognitive decline | Time points<br>□ | Prevalence % (95% CIs where available) |
| --- | --- | --- | --- | --- | --- | --- | --- | --- | --- |
| Buchanan 2015 [54] | Cross-sectional | United States | Chemotherapy = 288<br>Chemotherapy + hormone = 859 | 34 - 82 | FACT-Cog | High | Responses of cognitive complaints “sometimes”, “often”, or “always” | Mixed* | 74 Chemotherapy<br>78 Chemotherapy + hormone |
| Debes 2010 [50] | Cohort | Denmark | 75 | 29–59, 47.2 | Author-derived questions assessed on 7 point scale | High | Scores of 5–7 | T2 | Memory: 29<br>Concentration: 13<br>Mental fatigue: 33<br>Vigour: 43 |
| Ganz 2013 [55] | Cohort | United States | 189 | 51.8 | PAOFI | Moderate | Classified based on PAOFI score as high range versus normal/no complaints | Mixed* | Memory: 23<br>High level cognition: 19 |
| Hurria 2006 [45] | Prospective Longitudinal | United States | 45 | 65–84, 70 | Squire | Moderate | Unclear but based on score | T3 | 51 |
| Janelins 2017 [56] | Prospective Longitudinal | United States | 505 | 22–81, 53.4 | FACT-Cog | High | A 1/2 standard deviation representing a minimal clinically important difference | T2, T3 | T2: 45<br>T3: 37 |
| Jenkins 2006 [51] | Cohort | UK | 85 | 51.5 | Broadbent | High | Self-report of change | T2, T3 | T2:<br>Memory: 83<br>Concentration: 80<br>T3: |
|  |  |  |  |  |  |  |  |  | memory: 60<br>concentration: 45 |
| Koppelmans<br>2012 [11] | Cohort | Netherlands | 196 | 50-80,<br>64.1 | Basic self-report | High | Yes response to<br>question | T7 | More problems<br>remembering: 53<br>Forgetting (daily)<br>pursuits: 43<br>Word-finding<br>problems: 38 |
| Lange 2016<br>[15] | Cohort | France | 58 | 65 – 81,<br>70 | FACT-Cog | High | A difference of<br>more than 10%<br>between baseline<br>and T2 was<br>considered<br>clinically<br>significant | T2 | Perceived Cognitive<br>Impairment: 34<br>Perceived Cognitive<br>abilities: 49<br>(80 over 75 yrs) |
| Ng 2018<br>[52] | Prospective<br>Longitudinal | Singapore | 166 | 50.7 | FACT-Cog | High | A drop of 10.6<br>points in the global<br>score from<br>baseline. | T1, T3 | T1: 22<br>T3: 31 |
| Rey 2012<br>[57] | Cohort | France | 222 | 18-40, 37 | Telephone<br>Interview | High | Women who<br>reported memory<br>and/or attention<br>troubles “very<br>often” or “often” | T3, T4,<br>T5 | T3:37<br>T4: 37<br>T5: 42 |
| Schagen 1999 [53] | Cohort | Netherlands | 39 | 47.1 | Interview with questions scored on a Likert scale | Moderate | A score of 2 (moderate) or more was considered a distinct complaint about the cognitive functioning in the domain concerned. | T5 | Concentration problems: 31<br>Memory problems: 21<br>Thinking: 8<br>Language: 8 |
| Schmidt 2016 [58] | Cross-sectional | United States | 904 | 48.8 (across range of cancer types) | Based on Quality of Life in Adult Cancer Survivors (QLACS) with some amendment | High | An endorsement of the items was categorised as perceived cognitive decline. | Unclear | 58 |
| Shilling 2007 [59] | Cohort | UK | 85 | 51.7 | Interview | Moderate | An endorsement of the items | T2, T4 | Memory<br>T2: 83<br>T4: 60<br>Concentration:<br>T2: 78<br>T3: 45 |
| Tager 2010 [60] | Cohort | United States | 31 | 46.95–70.96, 60.7 | Participants asked to rate perceived memory abilities on a five-point Likert scale | Moderate | Ratings >2 were coded as cognitive problems | T2,T3 | T2: 43<br>T3: 46 |
| Van Dam<br>1998 [42] | RCT | Netherlands | FEC <sup>□</sup>:36<br>CTC<sup>#</sup>: 34 | FEC <sup>□</sup>:<br>48.1<br>CTC<sup>#</sup>:<br>45.5 | Cognitive<br>problems in<br>daily life<br>interview | High | An endorsement of<br>the items was<br>categorised as<br>perceived cognitive<br>decline. | T5 | Concentration:<br>High-dose:38<br>Standard-dose: 31<br>Memory<br>High-dose: 32<br>Standard-dose: 28<br>Thinking:<br>High-dose: 21<br>Standard-dose: 11<br>Language:<br>High-dose:12<br>Standard-dose: 11 |

\* Time-points measured in study cross time-points used in this review preventing accurate assimilation
□ T1 = During chemotherapy treatment, T2 = Just after cessation of treatment, T3 = 6 months after treatment cessation, T4 = 1 year after treatment cessation, T5 = 2-3 years after treatment cessation, T6 = 5 – 10 years after treatment cessation, T7= ≥10 years after treatment cessation
□ Fluorouracil, epirubicin, cyclophosphamide (standard dose)
<sup>#</sup> Four cycles of FEC followed by cyclophosphamide, thiotepa, and carboplatin (high dose)

### Methodological quality

A summary of the methodological quality assessment for the included articles is provided in Figure 2 and supplementary material (S7). Thirty-five (67%) studies were considered to be of high quality, fifteen (29%) of moderate quality and only two [43, 44] (4%) studies of low quality (see Tables 1, 2 and 3). The main domains of concern were participant sampling, sample size, condition measurement and statistical analysis. Of these, the outcomes of assessment of risk in the domains of sample size and condition measurement are considered to have the most impact on the review outcomes. However, use of a quality model in meta-analysis and evaluating the impact of sample size by meta-regression minimizes this impact.

**Figure 2:**
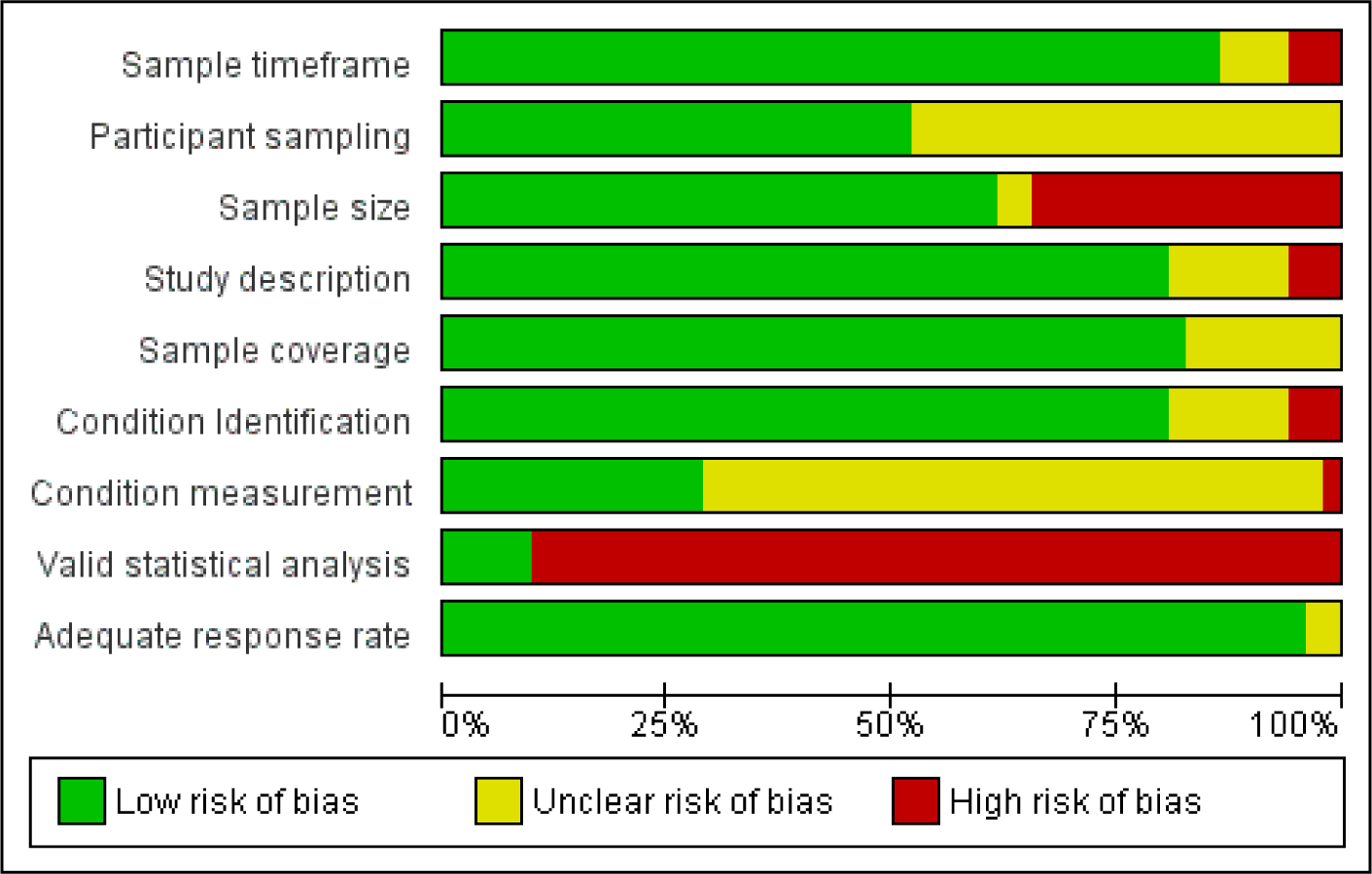
Risk of bias graph: review authors’ judgements about each methodological quality item presented as percentages across all included studies. Study number = 52.

**Table 2:** Summary of included studies employing short cognitive screening tools to assess cognitive impairment

| Study | Study Design | Location | Sample Size | Sample Age (Range, Mean) | Assessment Method | Methodological Quality | Definition of cognitive decline | Time point(s)<br>□ | Prevalence % (95% CIs where available) |
| --- | --- | --- | --- | --- | --- | --- | --- | --- | --- |
| Biglia 2012 [49] | Prospective Longitudinal | Italy | 40 | 38–65, 51 | MMSE | Moderate | Score under the mean | T2 | 31 |
| Brezden 2000 [25] | Cohort | Canada | 71 across two groups:<br>1) Currently receiving chemotherapy, and 2) one year after cessation | 34-70, 49<br>26-61, 46 | HSCS | High | Classification as mild, moderate or severe according to pattern of scores as described by test authors | T1, T4 | T1<br>mild: 13<br>moderate: 10<br>severe: 12/31 = 39<br>T4<br>mild: 20<br>moderate: 28<br>severe: 9/40 = 23 |
| Fan 2005 [26] | Cohort | Canada | 81 | 48 (median) | HSCS | High | Scores from each item on the test were subject to an algorithm which generated bands of impairment | T2, T4, T5 | T2<br>Mild: 35<br>Mod-Severe: 16<br>T4<br>Mild: 30<br>Mod-severe: 4<br>T5:<br>Mild: 21<br>Mod-severe: 4 |
| Fontes 2016 [40] | Prospective Longitudinal | Portugal | 475 | 54.7 (median) | MoCA | High | A MoCA score at least 2.0 standard deviations below age- and education-adjusted cut-offs | T3, T5 | T3: 7<br>T5: 8 |
| Ng 2018 [52] | Prospective Longitudinal | Singapore | 166 | 50.7 | Headminder computerized test | High | Reliable change index (RCI) score calculated to determine cognitive decline in each cognitive domain. A RCI score of lower than -1.5 was used as criteria for decline. | T1, T3 | T1: 6.1-21.6<br>T3: 1.2-14.6 |
| Pillai 2019 [43] | Prospective Longitudinal | India | 152 | 27 -72, 47 | MMSE | Low | MMSE score of $\leq 24$ (out of 30) | T2,T3,T4, T5 | T2: 0<br>T3: 0<br>T4: 0<br>T5: 0 |
| Prokasheva 2011 [61] | Cohort | Israel | 20 | 30-57, 49.3 | Doors and People Test | Moderate | Below 1SD as indicative of mild impairment and below 2SD for moderate impairment | T5 | 40 |
| Ramalho 2017 [62] | Cohort | Portugal | 418 | 27 -87 | MoCA | High | MoCA score values at least 1.5 standard deviations below age- and education-adjusted cut-offs | T3 | 8 (5.8,11) |
| Tchen 2003 [63] | Cohort | Canada | 110 | 27-60, 48 | HSCS | High | Based on standard test criteria | T1 | T1<br>Mild: 34<br>Moderate: 2<br>Severe: 14 |

□ T1 = During chemotherapy treatment, T2 = Just after cessation of treatment, T3 = 6 months after treatment cessation, T4 = 1 year after treatment cessation, T5 = 2-3 years after treatment cessation

**Table 3:** Summary of included studies employing objective neuropsychological testing to assess cognitive impairment

| **Study** | **Study Design** | **Location** | **Sample Size** | **Sample Age (Range, Mean)** | **Number of Tests in Battery** | **Methodological Quality** | **Definition of Cognitive Decline** | **Time point(s)**<br>□ | **Prevalence % (95% CIs where available)** |
| --- | --- | --- | --- | --- | --- | --- | --- | --- | --- |
| Andryszak 2018 [64] | Cohort | Poland | 31 | 52.4 | 1 | Moderate | Scores lower than 2 SD compared to control group averages | T1,T2 | T1:32<br>T2: 26 |
| Biglia 2012 [49] | Prospective Longitudinal | Italy | 40 | 38–65, 51 | 10 | Moderate | “Unsatisfying score” not defined in report | T2 | 15 |
| Collins 2009 [65] | Cohort | Canada | 53 | 50-65, 57.9 | 18 | High | SRB* scores of –2 or less on at least two tests | T2, T4 | T2: 34<br>T4: 11 |
| Collins 2013 [66] | Prospective longitudinal | Canada | 60 | 52.4 | 17 | High | Scores lower than 2 SD on at least two tests | T1<br>( measured at end of each cycle) | 30 (mean across all cycles) |
| Collins 2014 [67] | Cohort | Canada | 56 | 51.8 | 19 | High | SRB scores of –2 or less on at least two tests | T1<br>( measured at end of each cycle)<br>T4 | T1: 48<br>T4: 22 |
| Debess 2010 [50] | Cohort | Denmark | 75 | 29–59, 47.2 | 4 | High | Significant changes (between 5th and 95th percentile in controls) on at least two tests | T2 | 4 |
| Hermelink 2007 [68] | RCT | Germany | 101 | 48.6 | 12 | High | Reliable-change index (RCI) with a probability of error set at 10% | T1 | 27 |
| Hermelink | Cohort | Germany | 91 | 27.3–64.9, | 18 | High | Five or more scores | T2, T4 | T2: 6 |
| 2017 [69] |  |  |  | 52.3 |  |  | below 1.5 standard deviations and/or four below 2 standard deviations |  | T4: 18 |
| Hurria 2006 [46] | Prospective longitudinal | United States | 28 | 65–84, 71 | 13 | Moderate | Scores lower than 2 SD on at least two tests | T3 | 39 |
| Jansen 2011 [70] | Prospective longitudinal | United States | 71 | 30 – 65, 50.3 | 4 | High | Scores lower than 1 SD on at least two tests | T1, T2, T3 | T1:23<br>T2:52<br>T3:20 |
| Jenkins 2006 [51] | Cohort | UK | 85 | 51.5 | 7 | High | Reliable change index on at least two measurement | T2, T3 | T2: 20<br>T3: 18 |
| Jim 2009 [71] | Cohort | United States | 97 | 50 | 9 | High | Scores lower than 1.5 SD on at least two tests | T3 | 34 |
| Jung 2014 [72] | Cross sectional study | Korea | 32 | 31-61, 46 | 3 | High | Cutoff as 5/4 on Digit Span -DS F; 4/3 on Digit Span B; 28-30/25 on Controlled Oral Word Association- COWA) | T2 | DSF<br>mild: 32<br>moderate: 13<br>DSB<br>mild: 32<br>moderate: 42<br>COWA* (A)<br>mild: 3<br>moderate: 58<br>COWA (B)<br>mild: 52 |
| Kesler 2017 [73] | Cohort | United States | 31 | 34–65, 48.6 | 4 | Moderate | Scores lower than 1.5 SD on at least two tests or lower than 2SD on any one test. | T4 | 55 |
| Kreukels | Cohort | Netherland | 63 | 46.6 | 10 | High | Scores lower than 2 | T3/T4 | 33 |
| 2008 [74] |  | s |  |  |  |  | SD on at least three tests |  |  |
| Lange 2016 [15] | Cohort | France | 58 | 65 – 81, 70 | 8 | High | A significant change in a domain score | T2 | 64 |
| Mehlsen 2009 [75] | Cross-sectional | Denmark | 36 | 48.6 | 13 | High | Decline on at least 3 of the cognitive measures. | T2 | 29 |
| Mehnert 2007 [76] | RCT | Germany | 23 | 33–65, 53 | 18 | Moderate | Scores lower than 1.4 SD on at least four tests | T6 | 13 |
| Menning 2016 [77] | Cohort | Netherlands | 31 | 49.8 | 11 | High | Multivariate normative comparison comparing against distribution of scores in control group. | T3 | 16 |
| Reid-Arndt 2009 [78] | Cross sectional study | United States | 46 | 53.4 | 11 | High | Scores of below 1 SD | T2 | Executive Functioning: 22<br>Verbal Fluency: 41 |
| Reid-Arndt 2010 [79] | Prospective longitudinal | United States | 39 | 53.4 | 7 | High | Below 1SD as indicative of mild impairment and below 1.5 SD for moderate impairment | T3, T4 | WMS Log Mem:<br>T3: 6<br>T4: 33<br>RAVLT<br>T3:13<br>T4: 6<br>WMS Log Mem II<br>T3: 3<br>T4: 33<br>Trails A<br>T3: 3<br>T4: 9 |
|  |  |  |  |  |  |  |  |  | Trails B<br>T3: 15<br>T4: 24<br>SCW<br>T3: 3<br>T4: 3<br>COWA<br>T3: 0<br>T4: 9<br>Category Fluency:<br>T3: 0<br>T4: 16 |
| Ruzich 2007[80] | Prospective longitudinal | Australia | 35 | 30–66, 53 | 15 | Moderate | Scores below 1 SD on at least two tests | T1,T2,T3 | T1: 15<br>T2: 37<br>T3: 30 |
| Schagen 1999 [53] | Cohort | Netherlands | 39 | 47.1 | 14 | Moderate | Scores below 2 SD on at least three tests | T5 | 28 |
| Schagen 2002 [81] | Cross-sectional | Netherlands | Two groups of different chemotherapy regimes:<br>FEC<sup>□</sup>: 23<br>CMF<sup>a</sup>: 31 | 50.3 | 13 | High | Number of tests with scores below 2SD | T5 | FEC: 9<br>CMF: 13 |
| Schagen 2006 [82] | RCT | Netherlands | Two groups of different chemotherapy regimes:<br>FEC□:<br>39<br>CTC<sup>‡</sup>: 28 | 45 | 10 tests (not listed) | High | Scores below 2 SD on at least three tests | T3 | FEC: 10<br>CTC: 20 |
| Schrauwen<br>2020 [41] | Cohort | Belgium | 66 | 27 – 64, 46.7 | 5 | High | Standardized<br>difference score<br>exceeding -2.5 SD on<br>at least one test. | T2 | 24 |
| Shilling<br>2005 [83] | Cohort | UK | 50 | 51.1 | 8 | High | Reliable decline on<br>two or more tests | T2 | 34 |
| Stewart<br>2008 [84] | Cohort | Canada | 61 | 50-66, 57.5 | 18 | High | Two or more SRB<br>scores of -2.0 or less<br>on at least two tests | T2 | 31 |
| Stouten-Kemperma<br>n 2015<br>[85] | RCT | Netherland<br>s | Two groups of<br>different<br>chemoth<br>erapy<br>regimes:<br>Conventi<br>onal<br>Dose =24<br>High<br>Dose =<br>17 | 56.3- 59.8 | 8 | Moderate | Score larger than<br>2SDs below the mean<br>considered impaired<br>on a test. The fifth<br>percentile of the<br>overall impairment<br>score of healthy<br>control scores was<br>cutoff for impairment | T7 | Conventional Dose:<br>8<br>High Dose: 11 |
| Syarif<br>2019 [44] | Cross-sectional | Indonesia | 82 | 43 | 1 | Low | Not reported | Unclear | 87 (mild-serious<br>impairment) |
| Van Dam<br>1998 [42] | RCT | Netherland<br>s | FEC□:<br>36<br>CTC<sup>‡</sup>: 34 | FEC: 48.1<br>CTC: 45.5 | 13 | High | Scores below 2 SD<br>on at least three tests | T5 | FEC: 17<br>CTC: 32 |
| Van Dyk<br>2018 [86] | Cross-sectional | United<br>States | 20 | 47 | 17 | Moderate | Impaired by ICCTF<br>guidelines | T2 | 45 |
| Vearncombe 2009 [87] | Prospective longitudinal | Australia | 136 | 49.38 | 10 | High | Reliable change on at least 2 measures | T2 | 17 |
| Wefel 2010 [88] | Prospective longitudinal | United States | 42 | 33-65, 48,4 | 6 | High | Scores below 2SD on one test. | T2, T4 | T2:65<br>T4:61 |
| Wieneke 1995[89] | Cross sectional | United States | 28 | 28 – 54, 42 | 15 | Moderate | Score below 2 SD on at least one test classified as moderate impairment | T2-T4 (varies across period) | 75 |

□ T1 = During chemotherapy treatment, T2 = Just after cessation of treatment, T3 = 6 months after treatment cessation, T4 = 1 year after treatment cessation, T5 = 2-3 years after treatment cessation, T6 = 5 – 10 years after treatment cessation, T7 = $\geq 10$ years after treatment cessation
\*Standardised Regression Based
□ Fluorouracil, epirubicin, cyclophosphamide
<sup>a</sup> Cyclophosphamide, methotrexate, 5-fluorouracil
<sup>#</sup> Four cycles of FEC followed by cyclophosphamide, thiotepa, and carboplatin,

### Prevalence estimates based on self-report measures of cognitive impairment

It was considered that there was significant diversity in the studies that used self-report measures in terms of method of assessment of cognitive impairment, timing of assessment in relation to chemotherapy treatment and form of report (based on cognitive domain rather than overall impairment). Therefore, these results are presented narratively.

Fifteen studies (Table 1) examined prevalence of cognitive impairment after breast cancer treatment contributing 22 prevalence estimates across all study time-points with the exception of T6. There were a number of studies with large sample sizes in this group [54, 56, 58], however the median sample size for the group remained modest at 85 (range 31- 1147). Studies utilized a variety of methods to assess impairment which ranged from validated self-report instruments such as FACT-Cog or PAOFI, to semi-structured interviews or Likert responses to questions on cognitive function. Seven (47%) of studies used the former. When the studies employing validated tests were contrasted with those using non-validated methods median global impairment across the time points was 46% and 39.5% respectively, with no significant difference in prevalence rate between the two subgroups (two tailed T-test; *t* (6,5) = -1.2, *p* = 0.27).

Prevalence rates for global impairment varied from 21-83% (mean 44%) across all time- points. Noting the paucity of data at T6 and T7, no obvious downward trend in impairment across time is evident (Figure 3). Two studies in this sub-group focused on CICI in older women over the age of 65 [45], [15] reporting prevalence rates of 51% [45], and 34%. [15] Whilst, the former is clearly above the mean reported above of 44%, the value within Lange et al. 2016 [15] actually falls under the calculated average.

**Figure 3:**
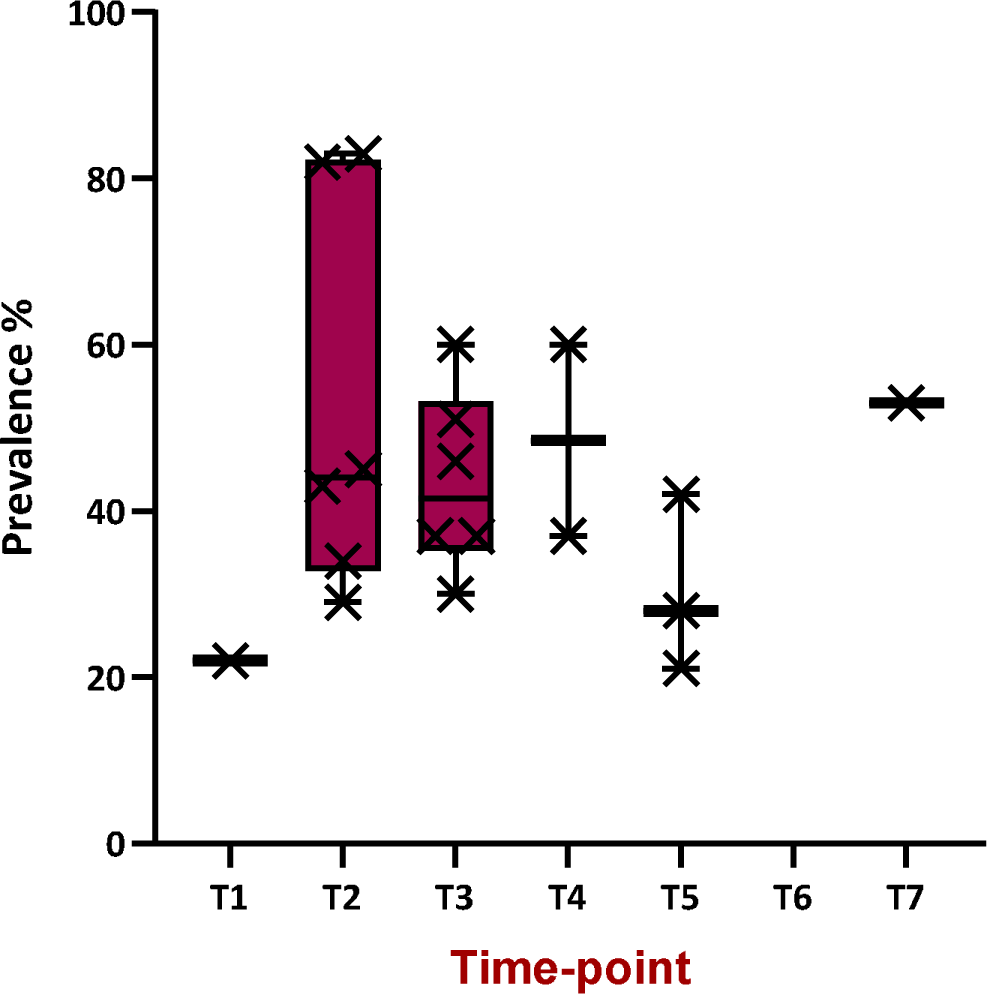
Boxplot of prevalence reported for cognitive impairment, via self-report methods, across the time-points included in the review (data from 12 studies in Table 1, 3 studies were excluded due to mixed or unclear timing of assessment [54, 55, 58])

### Prevalence estimates based on use of short cognitive screening tools to measure cognitive impairment

There was considerably less diversity in assessment method for cognitive impairment, and time points of measure for the studies that utilised short cognitive screening measures compared to those employing self-report. However, at each time point there were fewer than five studies. Study has suggested that at least 5 studies are needed to achieve statistical power from random effects meta-analyses that are superior to the individual contributing studies. [90] Studies within this group were therefore not subjected to meta-analytic techniques and are reported narratively.

Nine studies (Table 2) utilized short cognitive screening tools as part of this grouping, contributing 17 prevalence estimates. No studies performed longer-term follow up at the T6 and T7 time-points. Sample size ranged from 20-475 (mean =170). Five different validated methods of assessing impairment were employed (Table 1). With the exception of the Doors and People test these methods assess a range of cognitive domains expected to be impacted by chemotherapy administration. In two studies [26] [25] test outcomes were reported based on a banding of scores as mild, moderate or severe. For the purposes of assimilation, only prevalence estimates for moderate-severe impairment have been considered.

In contrast to studies that used self-report none of these studies specifically examined older populations. However, three of these studies [25, 43, 62] had a wider age range with patients above 65 years of age being included. This has not however led to a substantially increased mean participant age in comparison to the other studies.

Prevalence rates across all time-points ranged from 0-50. The mean prevalence was 16%. Exclusion of the four prevalence estimates of zero provided by Pilai et al. 2019 [43] leads to a range of 4%-50%, with mean prevalence of 21%. This action may be justified given the low quality rating assigned to this study. There was a general downward trend in prevalence across time, but range at each time-point (with the exception of T3) was large (Figure 4).

**Figure 4:**
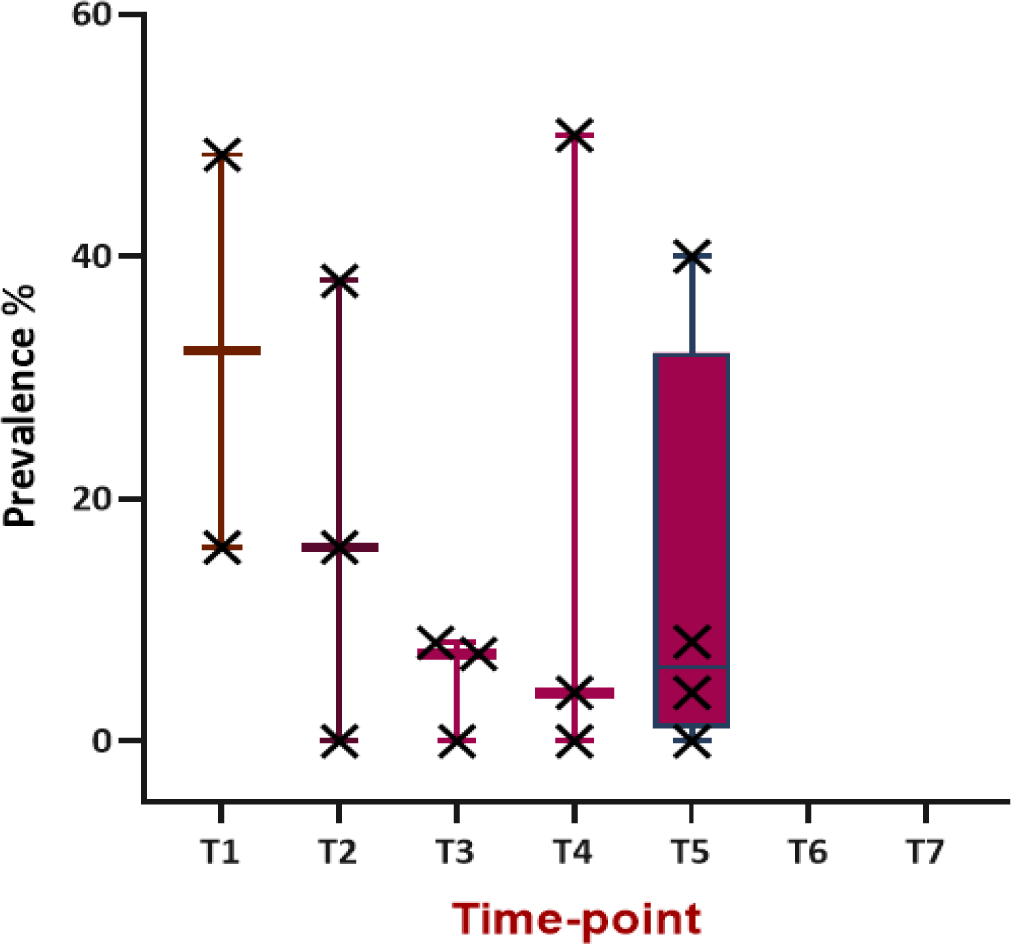
Boxplot of prevalence reported for cognitive impairment, via short cognitive screening methods, across the time-points included in the review. Ng et al. 2018 [52] was excluded from the calculation since a range was reported, and only considering moderate-severe classification of impairment from [26] and [25].

### Prevalence estimates based on use of objective neuropsychological testing to measure cognitive impairment

Thirty-five studies (Table 3) utilized objective neuropsychological tests to screen for cognitive impairment. These studies contributed 46 prevalence estimates. Sample size ranged from 20-136 (median = 53) having the lowest range and average of all the groups. Studies reported prevalence across all time points of interest, although only one estimate was reported at each of the longer-term follow up points (T6 and 7). Two studies in the group evaluated women over the age of 65. [15, 46] The neuropsychological tests used were all established, validated tests which when administered in a battery broadly encompass the major cognitive domains. Two studies only employed one test for cognitive assessment, [44, 64] and as such did not evaluate all cognitive domains.

There were differences between the studies in the criteria used to determine cognitive impairment. In general a diagnosis of impairment was made based on two factors being met: 1) the level of decline in score relative to some reference value, for example healthy control patients 2) that this decline occurred in a pre-determined number of tests. Fourteen (40%) of the studies used 2 standard deviation declines to guide level but there was heterogeneity even between these in the number of tests with declining scores required to satisfy the diagnostic criteria (see Table 3). Studies that considered a one SD decline as criteria for impairment, or were not clear on the level used, were considered less stringent (see meta-analysis). Three studies [72, 78, 79] reported test outcomes by cognitive domain or individual test, rather than global impairment rendering these data harder to assimilate with the other studies.

### Meta-analysis

#### Pooled Prevalence

It was considered that meta-analysis was appropriate for further investigation of prevalence estimates derived from studies utilizing objective neuropsychological tests. Thirty-seven prevalence estimates from 27 articles were included in the meta-analysis, grouped based on time point of prevalence measure (from T1-T5). Only one study contributed data at each of T6 [76], and T7 [85] and consequently these time points and studies were not included. Three studies were excluded from entry since prevalence was reported based on cognitive domain or test type [72, 78, 79]. A further three studies were excluded since the timing of measure was unclear [44], or was performed across multiple time points as defined for this review. [74, 89] In two studies, prevalence estimates were reported based on chemotherapy regime. [81] [82] In order to ensure comparability with the other included studies only estimates for the standard dose chemotherapy regime from Schagen et al. 2006 [82] were included in the meta-analysis. Prevalence estimates reported by chemotherapy regimen in Schagen et al. 2002 [81] were pooled.

The overall random-effects pooled prevalence (95% CIs) of cognitive impairment from time points T1-T5 respectively was 34% (24-44), 30% (20-40), 23% (16-31), 31% (14-50%), and 21% (15-28) (Figure 5). Figure 6 synthesises these findings narratively across the time- points with the inclusion of the single individual study estimate contributed from [76] and [85] at T6 and T7. There is a general downward trend in prevalence across time, although substantial imprecision in the effect at T4 with a slightly increased prevalence than might be expected if observing the trend.

**Figure 5:**
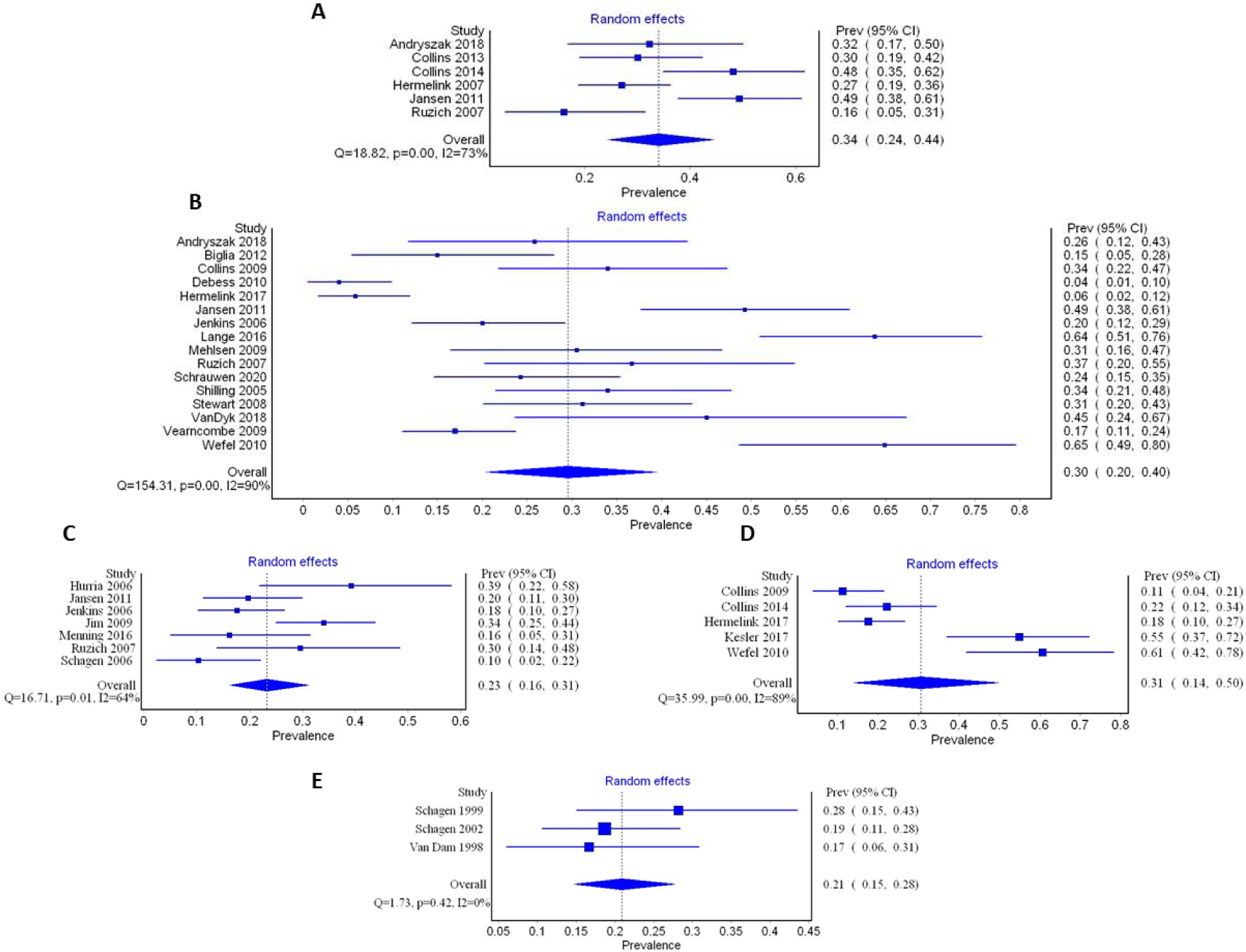
Forest plots of prevalence reported in studies utilising neuropsychological tests to diagnose cognitive impairment following chemotherapy treatment for breast cancer. A) Cognitive assessment undertaken during chemotherapy treatment (T1), B) Cognitive assessment undertaken just after cessation of chemotherapy treatment (T2), C) Cognitive assessment undertaken 6 months after cessation of chemotherapy treatment (T3), D) Cognitive assessment undertaken 1 year after cessation of chemotherapy treatment (T4), E) Cognitive assessment undertaken 2-3 years after cessation of chemotherapy treatment (T5).

**Figure 6:**
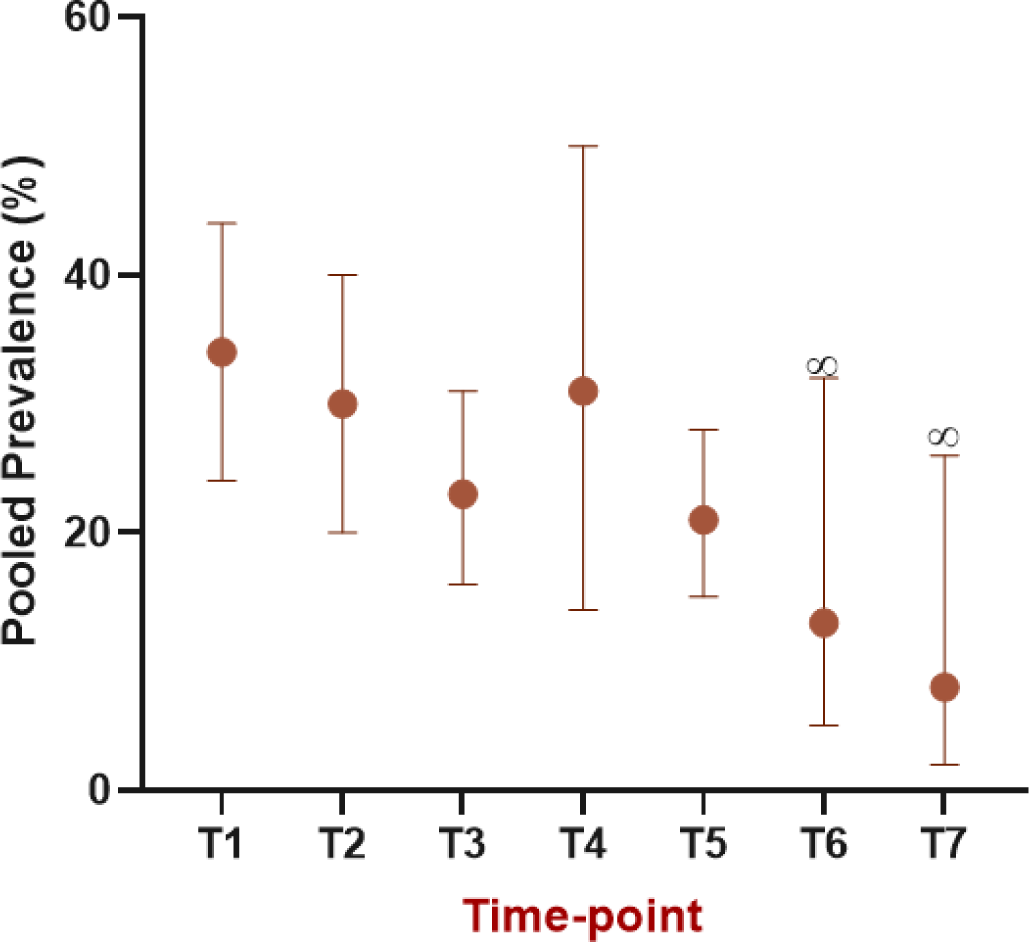
Prevalence reported for cognitive impairment, via neuropsychological test methods, across the time-points included in the review. Data represent pooled prevalence 32 estimates derived from meta-analysis, with 95% CIs. ∞ - values are not pooled figures but represent individual values from one study

#### Heterogeneity and Publication Bias

Heterogeneity as determined by the *I*^2^ statistic was universally high with the exception of T5 (where there were limited estimates) at 73%, 90%, 64%, 89% and 0 (T1-5 respectively).

Sensitivity analyses performed included removal of each study in turn, analysing untransformed data, considering the effect of the quality score and using a fixed effects model. In general, results were similar after these analyses (supplementary material S8). However, removal of Jansen et al. 2011 [70] at T1 had a significant effect reducing *I*^2^ by 10% to 63% (potentially as a result of the impairment criteria used as described below). It is noteworthy that removal of the Andrysak 2018 study, [64] which only employed one cognitive test, did not impact the heterogeneity estimates, nor did removal of the studies with cross-sectional designs. [75, 86] In order to explain the heterogeneity in the pooled effects sizes the influence of stringency of criteria for diagnosing cognitive impairment was explored via subgroup analysis. Only prevalence reports from T2 and T3 were subjected to subgroup analyses based on the number of studies contributing data at these time-points, and the number in each analysis when split into sub-groups. At these time-points, four studies were considered to have used less stringent test methods. [70, 75, 80, 88] Heterogeneity remained high after sub-group analysis with the high stringency sub-group having a pooled prevalence of 25% (95% CIs: 16-35), *I*^2^= 90% at T2, and 23% (95% CIs: 13-34), *I*^2^= 74% at T3 (Figure 7).

**Figure 7:**
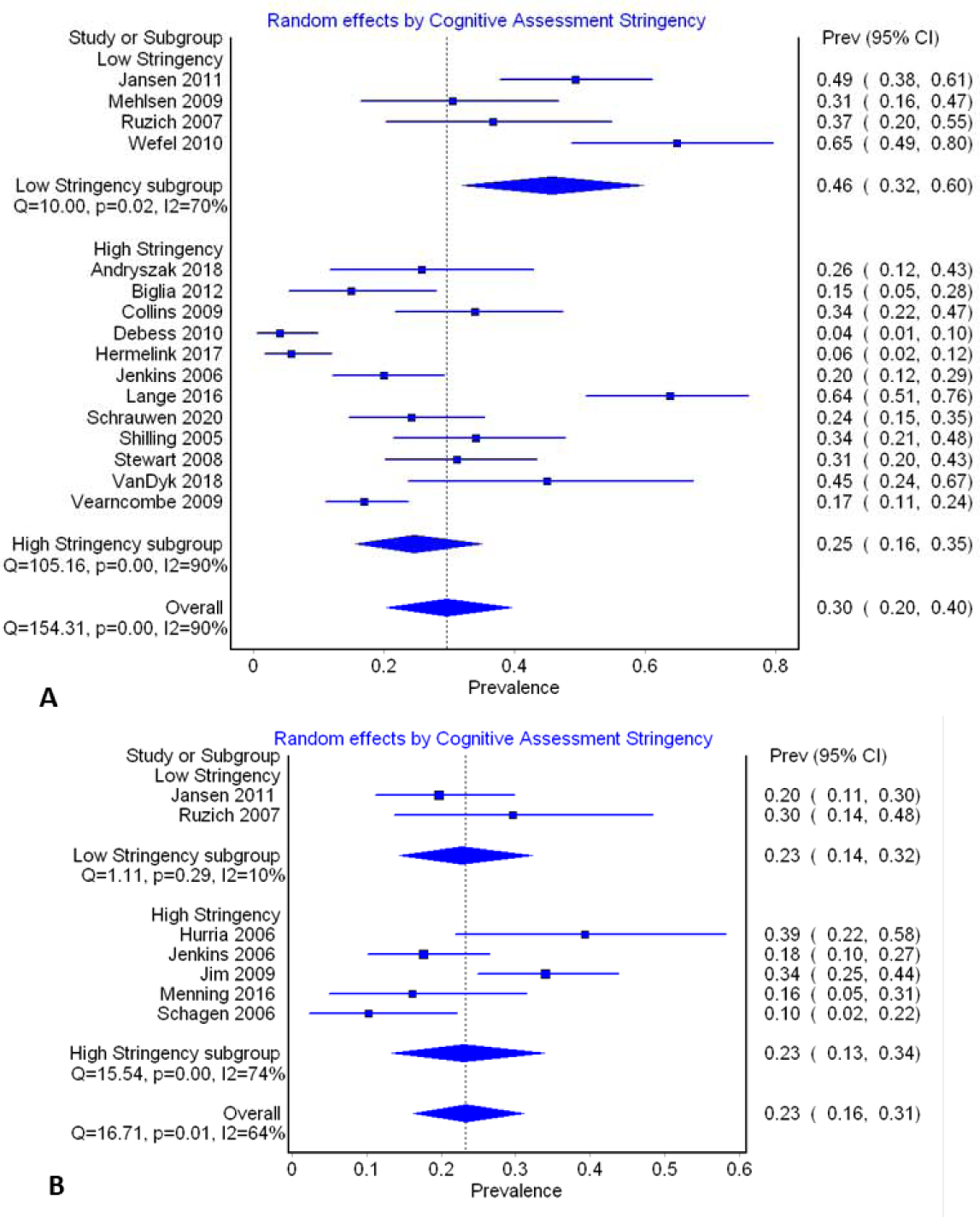
Forest plots of prevalence sub-grouped by stringency of cognitive assessment. A) Cognitive assessment undertaken just after cessation of chemotherapy treatment (T2), B) Cognitive assessment undertaken 6 months after cessation of chemotherapy treatment (T3)

Meta-regression was performed in an attempt to account for the remaining heterogeneity. The results of five individual meta-regression analyses performed at T2 based on 1) the continuous variables of age, methodological quality and sample size, and 2) categorical variables of cognitive impairment criteria stringency and geographical location (3 subsets; Europe (reference group), North America, Asia-Pacific) are included in Table 4. Cognitive impairment stringency explained some of the variance in prevalence, accounting for 16% of the heterogeneity at T2 (*P* = 0.04). Sample size also accounted for 16% of the heterogeneity (*P* = 0.04) with increased sample size tending to result in a lower prevalence estimate. The remainder of the covariates could not account for the variance observed.

**Table 4:**
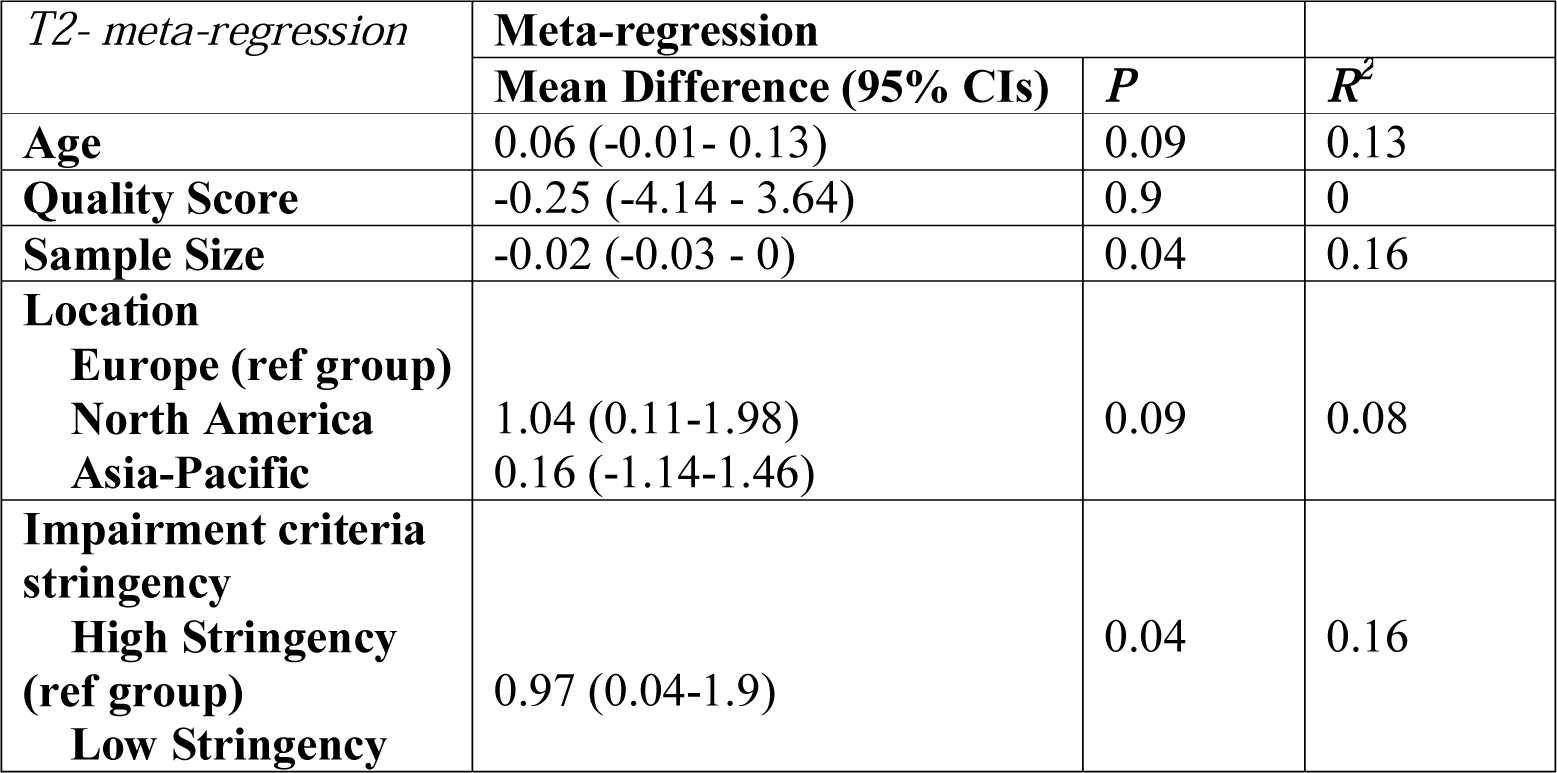
Results of four individual meta-regression analyses at T2 based on age, quality score, location and cognitive impairment criterion stringency

Meta-regression was then used to examine the relationship between sample size, cognitive impairment test stringency, and effect size. A test of this model yielded a Q-value of 6.99 with 2 degrees of freedom and corresponding p-value of 0.03. The test for heterogeneity yields a Q-value of 74.7 and a corresponding p-value of 0, implying that the variation of observed effects about the regression line falls within the range that cannot be explained by sampling error alone. The model is able to explain some 23% of the variance in true effects.

The doi plots for publication bias showed some asymmetry implying the presence of bias. The LF asymmetry index confirmed there was minor asymmetry at T2, and T5 and major asymmetry at T4. There was no evidence of publication bias at T1 and T3 (see supplementary material S9).

### GRADE certainty assessment and results

The evidence presented across all the studies grouped by cognitive assessment methods was assessed using the GRADE (Grading of Recommendations Assessment, Development and Evaluation) approach (GRADEpro GDT: GRADEpro Guideline Development Tool [Software]). Results are presented in the Summary of Findings Table. The certainty of evidence was graded as very low for all three-assessment types. Inconsistency was rated as serious for all groups of studies based on the wide variation in estimates and high calculated heterogeneity (neuropsychological testing). Imprecision was similarly rated as serious for all outcomes as a result of the wide confidence intervals. Publication bias was strongly suspected for the study group that used neuropsychological tests based on the doi plots and LFK index. Due to the method of synthesis used for the self-report and short cognitive screening tool groups, publication bias was not able to be assessed and was therefore rated as undetected.

## Discussion

This is the first systematic review on prevalence of CICI to have included and compared the three commonly used assessment modalities of self-report, short cognitive screen and objective neuropsychological tests. In contrast to the only other published systematic review of prevalence in CICI [3], meta-analytic techniques were applied to allow reporting of pooled effects, and studies which performed cognitive assessment beyond the breast cancer treatment period were also included. Prevalence estimates derived ranged from 0-83% across all time- points and all cognitive test methods. This crude figure is highly comparable with the commonly cited range of 12-82% based on the narrative review of Janelsins et al. 2014 [14]. The current review allows greater examination of variability in these prevalence rates, through examination of the influence of factors such as test method, time since treatment and age on rates.

### Impact of assessment tool choice on prevalence rates

There are striking differences in the average and maximum prevalence values reported based on method of cognitive assessment. Self-report of cognitive impairment yielded the highest values. There is substantially less variability between values obtained by short cognitive screen and neuropsychological test batteries. This difference in outcomes is to be expected for a number of reasons: 1) patient mood has been shown to have a relatively greater impact on subjective reports of impairment than actual cognitive performance. For example, aspects of mood such as anxiety, depression, poor quality of life and fatigue have all been shown to correlate with self-reported cognition complaints. [49, 91–93]; 2) expectations about treatment might influence self-report of cognitive symptoms. Study has shown that patients who were aware of the potential for cognitive side effects were more likely to report cognitive function issues than those who were unaware of the possibility; [94] 3) patients may have been functioning above the normal range of cognitive function prior to treatment, so whilst they experience a decline, their cognitive performance falls within the normal range; [95, 96] 4) the battery of objective tests used are ineffective at diagnosing subtle impairment [95, 96], this may especially be of concern with the use of short cognitive screening tools [97]; 5) objective tests are ecologically invalid by failing to represent real-life situations where patients may experience the impairment. [95, 96] Finally, based on the studies included in this review it could be argued that some of the assessment criteria are relatively crude, for example the endorsement of an item with no grading of response [11, 58, 59] would likely lead to substantially higher reporting of issues, which may not actually differ from normal age-related memory loss experienced by healthy individuals [11, 58, 59]. In fact a cursory glance at the prevalence reports in these studies shows that reported prevalences are higher than the mean for the group. This perhaps highlights a future need to only use validated self-report instruments for cognitive assessment.

In spite of the reduced variability observed with objective tests, there was still significant statistical heterogeneity observed on meta-analysis. Approximately 16% of this heterogeneity could be explained by the use of less stringent test criteria for diagnosing impairment.

Additionally, pooled prevalence was noticeably altered when subgroups based on test stringency were analysed. This finding provides strong evidence of the need for researchers to agree to, and use standardised impairment criteria, in order to make findings comparable, reproducible, and ultimately useful to practitioners.

### Impact of time since treatment on prevalence rates

Prevalence rates ascertained by short screening methods or larger batteries of tests showed a general downward trend over time, with a peak rate occurring during chemotherapy treatment. This finding is supported by pre-clinical and imaging studies in CICI, which demonstrate that, over time, partial resolution of the brain structural and functional impairments occurs. [22, 23] However, it does contrast with the trend in prevalence determined by Dijkshoorn et al. 2021 [3] which only included studies using objective tests, and found elevated (although perhaps not significantly so) prevalence rates at greater than one year after treatment, compared to just after treatment with chemotherapy. Notably however, actual prevalence values calculated in the current review for objective tests are remarkably similar to those in Dijkshoorn et al. 2021 [3]. Rates of 25%, 14 % and 27% were reported in that review at time-points of just after treatment, up to 1 year following, and greater than 1 year after treatment respectively. [3] This corresponds with 30%, 23% and 31% at equivalent time points in the current review. This similarity in rates is in spite of the differences in methodology employed in the two reviews with the current review assimilating rates via meta-analytical techniques, and the previous review only including longitudinal studies such that impairment was based on a decline in cognitive function from baseline measures.

Conversely, the current review illustrates that prevalence rates as determined by self-report remain high (often above 40%), and show little evidence of decline over time. The previously discussed suggestions explaining the general elevation in self-reported cognitive decline over measured impairment may also be at play here. Furthermore, at late follow-up stages patients will have likely returned to normal daily-life activities, such as work, and may have a greater perception of impairments as they return to navigating everyday tasks.

Whilst fewer studies have evaluated longer-term follow-up points, greater than 5 years following treatment cessation, there is evidence from both self-report and objective testing that some individuals are cognitively impaired at these time-points. This is an important finding for survivorship care and deserves dedicated research attention to fully elucidate impact.

### Impact of age on prevalence rates

There is recognition of the potential for increased significance of chemobrain in older adults with cancer due to higher levels of pre-existing cognitive impairment in this age group. [98] Furthermore, older adults may be more susceptible to cancer treatment toxicity leading to heightened symptoms. [15] Whilst it might therefore be predicted that CICI prevalence increases with age, the review did not find enough evidence to either support or refute this.

When meta-regression was applied, age in general was shown to have no effect on the high heterogeneity seen at T2. This lack of effect is unsurprising given that the studies included in the meta-regression had similar mean ages of the sample population, with relatively narrow ranges. Furthermore, for objective testing cognitive score results are usually compared against age-matched controls or some other age-based correction is applied, hence age is implicitly accounted for in experimental design.

Only three studies specifically focused on patients over the age of 65 [15, 45, 46]. Two of these studies [45] [15] used self-report methods, with one reporting prevalence rates below the mean for the group [45], and the other reporting rates high than the mean. [15] The third study [46] utilized objective tests and reported a prevalence value higher than the reminder of the T3 subgroup. However, it also has the widest confidence intervals of the group suggesting the estimate to be imprecise. The study also used a low sample size, which may have influenced the estimate obtained. Noteworthy, is that the cohort study of Lange et al. 2016 also used cognitive tests, with a diagnostic criteria categorized in this review as high stringency, and reported a high prevalence of 64% at T2. Moreover, confidence intervals were of similar width to other studies within the group. [15] This finding is perhaps the most suggestive of an effect of age on CICI prevalence and would be good grounds for dedicating future research effort to this question.

### Strength of Evidence on CICI prevalence

In order to account for any effect that low quality studies might have had on the review findings, the meta-analyses incorporated a quality model, and a meta-regression using quality score as a variable was performed. These tools indicated that quality had minimal effect on prevalence rates or variances in the pooled estimates. As a result, there can be confidence that study quality has not affected review outcomes, at least for those studies that utilized objective cognitive assessment tools. Whilst, it is harder to make a formal determination of the impact of quality on the prevalence reports from studies using self-report and short cognitive screening tools, there is little to indicate that the results may have been unduly influenced by poor quality studies.

In all of the meta-analyses performed, with the exception of T5 where there were limited studies, moderate to high statistical heterogeneity was found. Sensitivity testing failed to account for any meaningful heterogeneity, and the subgroup analyses failed to reduce *I*^2^ values substantially. Meta-regression did reveal an effect of cognitive impairment test stringency and sample size on variance. These factors accounted for 23 percent of the variance in effect seen at T2 in a combined model. The remainder of the heterogeneity could possibly be accounted for by methodological diversity due to the differing test batteries used, and further considerations relating to defining impairment levels that were not considered by the sub-group analysis. Additionally, clinical heterogeneity may have arisen as a result of treatments administered, patient menopausal status, or race.

There was evidence of publication bias at a number of time-points included in the meta-analysis, which lowers certainty in the findings from this analysis. However, there are a few considerations regarding assessment of publication bias in prevalence studies. Firstly, there is no specific guidance on assessment of publication bias when proportional data are involved. The issue being that funnel plots may be imprecise at the extremes of a proportion, [99] and this may similarly apply to doi plots. It could be envisaged that the risk of publication bias should be lower in studies of prevalence since there is no favourable hypothesized outcome that may influence decision to publish. However, in the current review it is worth noting that in all of the studies the study objective was not solely around obtaining prevalence data on CICI. Taking this into consideration the possibility of publication bias likely exists, and was evidenced in this study.

Certainty of the evidence contributing to the findings of this review was assessed using the GRADE approach. It was determined that there was low certainty in the body of evidence contributing to the review. However, there is a lack of formal guidance from the GRADE working group on use of the methodology for reviews of prevalence, and the guidance criteria are not always applicable to these types of data. In spite of the lack of standardization, based on the assessment conclusions should be made tentatively.

### Study Limitations

The review does have a number of limitations. First, due to the methodological diversity of the included studies, it was only possible to perform meta-analytic techniques on the studies that used neuropsychological tests. As a result, study weight has not been accounted for in the summary prevalence figures reported arising from self-report and short cognitive screening methods. This necessitates a need for extra circumspection around these values. Furthermore, even when meta-analytical techniques were used there was a resultant high statistical heterogeneity. Subgroup analyses and meta-regressions failed to account for most of this heterogeneity leaving the source(s) unknown, although some suggestions have been provided.

Second, due to the nature of the included studies with many having a repeated measures design it was not possible to use meta-analytical techniques to statistically compare prevalence across time-points; this being an aim of this review. Furthermore, in the current study using the All Time Points Meta-analysis approach assumption of independence between time-points was violated since some subjects contributed data at more than one time- point, whilst others only contribute at one time-point. This can lead to an ecological fallacy with a trend being seen on aggregate, that is not present at an individual level. [36] As a result there can only be low confidence in the direction of trends across time reported. Alternate approaches may be preferred when trends across time are of interest; for example, the use of a trend meta-analysis which uses regression modelling, or a change-in time meta-analysis where the primary study data is used to calculate differences between successive time-points or compared to baseline. [36] These alternate methods were not employed in this review since they require certain data to be provided in the primary studies such as a slope estimate for trend, or assume a longitudinal study design.

A few points regarding the conclusions that can be taken from this review are pertinent. The focus of the review was on estimating prevalence of CICI. However, as previously described cognition deficits may be caused by a range of factors associated with the cancer experience [14]. These may include the cancer itself and associated psychological distress and fatigue, or other administered treatments such as hormonal therapies. Additionally, studies eligible for inclusion in the review were not restricted to those with a longitudinal design, thus allowing for the contrast of findings with a baseline cognitive score. Based on these two points, it is impossible to say whether the cognitive deficits reported were wholly the result of chemotherapy, as opposed to a combination of cancer and multiple treatment-related effects. This concern is academic. From the perspective of patients and health-care providers it is the nature of cognitive decline that is important, not the origin of it. Since, these combinations of factors are inherent, and common to most breast cancer patients, it is considered that the review findings are generalizable.

### Conclusions

This review is the first systematic comparison of prevalence rates for cognitive impairment in breast cancer patient that has considered all methods used to ascertain impairment, and evaluates long-term prevalence. Mean prevalence rates for CICI across all time-points were 44% using self-report and 6% using short cognitive screening. Pooled prevalence rates of between 21-34% were derived for impairment diagnosed by objective neuropsychological tests. For all three assessment modalities the GRADE certainty in the evidence was rated as very low. Results therefore need to be interpreted with caution since the actual prevalence may be substantially different from this estimate.

#### Recommendations for practice

The findings of this review suggest that cognitive impairment may impact up to 1 in 3 patients at a level that is clinically significant. This impairment also extends beyond the treatment period and is often still apparent 2-3 years post treatment cessation. Health practitioners should consider the potential for cognitive impairment in breast cancer survivors, and guide them and their families towards sources of information and support. They may also like to discuss with patients some of the minimally invasive, non-therapeutic interventions that might ameliorate symptoms, for example cognitive training or physical activity [32].

#### Recommendations for research

The uncertainty around the true prevalence of cognitive impairment following cancer treatment leads to under-consideration of the condition, and consequently a lack of support being available for survivors. This leads to reduced patient quality of life through less meaningful engagement with work, family and daily activities, as well as economic impacts for both the individual and society. This review has highlighted a number of considerations for future research to provide greater certainty in the effect estimates. This includes increasing study sample sizes, utilising standardised cognitive test methods and batteries, and applying accepted, clinically relevant criteria for diagnosing impairment. With the increasing number of older cancer survivors, there is also a need for targeted research investigating this group. Acquiring accurate prevalence data will better inform survivorship strategies by allowing appropriate delivery of targeted resources and services, and informed resource allocation for this effort. Access to accurate prevalence estimates will also inform, and perhaps encourage, the development of evidence-based practice guidelines for CICI.

## Declaration of Competing Interest

The authors declare that they have no known competing financial interests or personal relationships that could have appeared to influence the work reported in this paper

## Funding

This research did not receive any specific grant from funding agencies in the public, commercial, or not-for-profit sectors. ALW was supported by a NHMRC Peter Doherty Biomedical Research Fellowship (APP1140072). RPG was supported by an Australian Government Research Training Program Scholarship.

## Data Availability

Data are included in manuscript and supporting files

## Notes

### Competing Interest Statement

The authors have declared no competing interest.

### Clinical Trial

Meta-analysis of published studies

### Author Declarations

Ethical approval not needed since meta-analysis

