## Supplemental Files for "Prevalence of cognitive impairment following chemotherapy treatment for breast cancer: A systematic review and meta-analysis"

### **Supplementary Material**

### **S1: Search Strategies (searches were conducted on 29th December 2020)**

MEDLINE via PubMed search strategy

| 1 | neoplasm[MeSH Terms] | "neoplasms"[MeSH Terms] |
| --- | --- | --- |
| 2 | cancer*[Title/Abstract] OR neoplas*[Title/Abstract] OR patient[Title/Abstract] OR survivor[Title/Abstract] | "cancer*"[Title/Abstract] OR "neoplas*"[Title/Abstract] OR "patient"[Title/Abstract] OR "survivor"[Title/Abstract] |
| 3 | #1 or #2 | "neoplasms"[MeSH Terms] OR "cancer*"[Title/Abstract] OR "neoplas*"[Title/Abstract] OR "patient"[Title/Abstract] OR "survivor"[Title/Abstract] |
| 4 | chemo*[Title/Abstract] | "chemo*"[Title/Abstract] |
| 5 | ("antineoplastic combined chemotherapy protocols") [MeSH Terms] | "antineoplastic combined chemotherapy protocols"[MeSH Terms] |
| 6 | # 4 or #5 | 4[UID] OR "antineoplastic combined chemotherapy protocols"[All Fields] |
| 7 | chemobrain[Title/Abstract] OR chemofog[Title/Abstract] OR "chemotherapy induced cogn*"[Title/Abstract] OR CICI[Title/Abstract] OR " chemotherapy related cognitive changes"[Title/Abstract] OR CRCI[Title/Abstract] OR neuropsychol*[Title/Abstract] | "chemobrain"[Title/Abstract] OR "chemofog"[Title/Abstract] OR "chemotherapy induced cogn*"[Title/Abstract] OR "CICI"[Title/Abstract] OR "chemotherapy related cognitive changes"[Title/Abstract] OR "CRCI"[Title/Abstract] OR "neuropsychol*"[Title/Abstract] |
| 8 | "cognitive dysfunction" or "cognition disorders" or " memory disorders" or "language disorders" or "neuropsychology" or "neuropsychological tests"[MeSH Terms] | "cognitive dysfunction"[All Fields] OR "cognition disorders"[All Fields] OR "memory disorders"[All Fields] OR "language disorders"[All Fields] OR "neuropsychology"[All Fields] OR "neuropsychological tests"[MeSH Terms] |
| 9 | # 7 or #8 | 7[UID] OR "cognitive dysfunction"[All Fields] OR "cognition disorders"[All Fields] OR "memory disorders"[All Fields] OR "language disorders"[All Fields] OR "neuropsychology"[All Fields] OR "neuropsychological tests"[MeSH Terms] |
| 10 | #3 and #6 and #9 | ("neoplasms"[MeSH Terms] OR ("cancer*"[Title/Abstract] OR "neoplas*"[Title/Abstract] OR "patient"[Title/Abstract] OR "survivor"[Title/Abstract])) AND (4[UID] OR "antineoplastic combined chemotherapy protocols"[All Fields]) AND (7[UID] OR ("cognitive dysfunction"[All Fields] OR "cognition disorders"[All Fields] OR "memory disorders"[All Fields] OR "language disorders"[All Fields] OR "neuropsychology"[All Fields] OR "neuropsychological tests"[MeSH Terms])) |

##### 1. SCOPUS search strategy

TITLE-ABS-KEY ( cancer*OR neoplas*orAND and patient OR survivor) AND TITLE-ABS-KEY ( chemo*OR "antineoplastic agents"OR "cancer treatment") AND TITLE-ABS-KEY ( chemobrain OR chemofog OR "chemotherapy induced cogn*"OR cici OR " chemotherapy related cognitive changes" OR crci OR neuropsychol*) AND ( LIMIT-TO ( DOCTYPE , "ar") ) AND ( LIMIT-TO ( LANGUAGE , "English") ) AND ( LIMIT-TO ( EXACTKEYWORD , "Human") )

##### 2. CINAHL search strategy

| S1 | cancer*[Title/Abstract] OR neoplas*[Title/Abstract] OR patient[Title/Abstract] OR survivor[Title/Abstract] OR neoplasm[MeSH Terms] |
| --- | --- |
| S2 | ("antineoplastic combined chemotherapy protocols") [MeSH Terms] OR chemo*[Title/Abstract] |
| S3 | "cognitive dysfunction" or "cognition disorders" or " memory disorders" or "language disorders" or "neuropsychology" or "neuropsychological tests"[MeSH Terms] OR chemobrain[Title/Abstract] OR chemofog[Title/Abstract] OR "chemotherapy induced cogn*"[Title/Abstract] OR CICI[Title/Abstract] OR " chemotherapy related cognitive changes"[Title/Abstract] OR CRCI[Title/Abstract] OR neuropsychol*[Title/Abstract] |
| S4 | #S1 AND #s2 AND #s3 |
| S5 | #S1 AND #s2 AND #s3 |

##### 3. PsycINFO search strategy

1	(cancer*OR neoplas*OR patient or survivor).mp. [mp=title, abstract, heading word, table of	contents, key concepts, original title, tests & measures, mesh]

2	exp Neoplasms/

3	1 or 2

4	exp Chemotherapy/

5	chemo*.mp. [mp=title, abstract, heading word, table of contents, key concepts, original title, tests & measures, mesh]

6	4 or 5

7	(chemobrain or chemofog or "chemotherapy induced cogn*" or CICI or " chemotherapy related cognitive changes" or CRCI or neuropsychol*).mp. [mp=title, abstract, heading word, table of contents, key concepts, original title, tests & measures, mesh]

8	("cognitive dysfunction" or "cognition disorders" or " memory disorders" or "language disorders" or "neuropsychology" or "neuropsychological tests").mp. [mp=title, abstract, heading word, table of contents, key concepts, original title, tests & measures, mesh]

9	exp Cognitive Impairment/ or exp Cognitive Ability/

10	7 or 8 or 9

11	3 and 6 and 10

##### S2: Data Extraction Template

**General information**

Title

Title of paper / abstract / report that data are extracted from

Country in which the study conducted

United States

UK

Canada

Australia

Other

Notes

**Characteristics of included studies**

Methods

Aim of study

Study design

Randomised controlled trial

Non-randomised experimental study

Cohort study

Cross sectional study

Case control study

Systematic review

Qualitative research

Prevalence study

Case series

Case report

Diagnostic test accuracy study

Clinical prediction rule

Economic evaluation

Text and opinion

Other

Population description**-**Age/setting of study

Stage of treatment

Currently undergoing treatment

In remission

Treatment**-**Chemotherapy and/or endocrine therapy.

Identified confounds

Total number of participants

Timepoint of cognitive report**-**Include this in relation to diagnosis +/- remission as relevant to study

Method of cognitive assessment

Self-report

Objective Neuropsychological testing

Imaging

Notes on specifics of cognitive assessment

Author's criteria for determining that cognition is impaired

Prevalence report for chemobrain (% and dispersion)

##### S3: Critical Appraisal Checklist. For each of the nine domains a score was assigned from 0-2, with 0 representing that the criteria had not been met, 1 being unclear or insufficient information, and 2 for criteria met. The domain scores were totaled (maximum obtainable 18) and the percentage of the maximum score computed for each study.

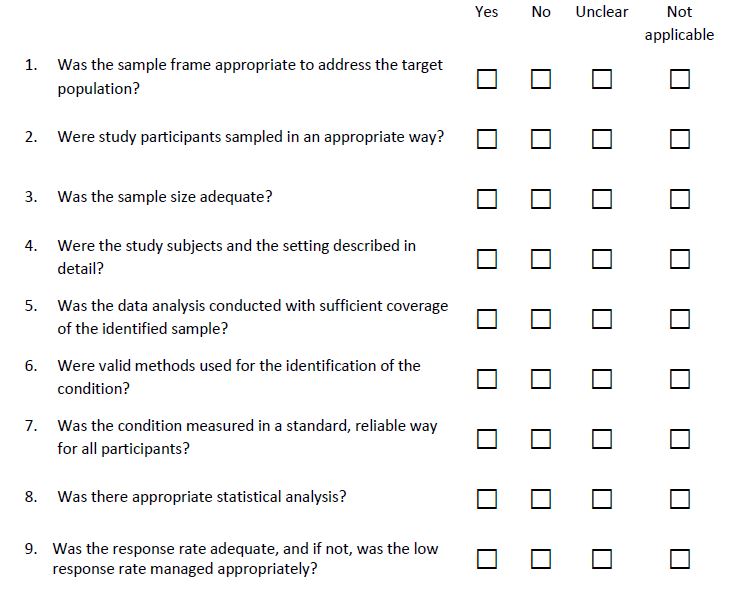

##### S4. List of excluded studies with reasons

| Study | Full Citation | Reason for Exclusion |
| --- | --- | --- |
| Ahles 2002 | Ahles TA, Saykin AJ, Furstenberg CT, Cole B, Mott LA, Skalla K, et al. Neuropsychologic impact of standard-dose systemic chemotherapy in long-term survivors of breast cancer and lymphoma. Journal of Clinical Oncology. 2002;20(2):485-93. | Outcomes not split based on cancer type |
| Anstey 2015 | Anstey KJ, Sargent-Cox K, Cherbuin N, Sachdev PS. Self-Reported History of Chemotherapy and Cognitive Decline in Adults Aged 60 and Older: The PATH Through Life Project. J Gerontol A Biol Sci Med Sci. 2015;70(6):729-35. | No prevalence data reported |
| Cheung 2012 | Cheung YT, Shwe M, Chui WK, Chay WY, Ang SF, Dent RA, et al. Effects of chemotherapy and psychosocial distress on perceived cognitive disturbances in Asian breast cancer patients. Annals of Pharmacotherapy. 2012;46(12):1645-55. | No prevalence data reported |
| deRuiter 2011 | de Ruiter MB, Reneman L, Boogerd W, Veltman DJ, van Dam FS, Nederveen AJ, et al. Cerebral hyporesponsiveness and cognitive impairment 10 years after chemotherapy for breast cancer. Human Brain Mapping. 2011;32(8):1206-19. | Multifaceted intervention with some patients receiving stem cell support |
| Donovan 2005 | Donovan KA, Small BJ, Andrykowski MA, Schmitt FA, Munster P, Jacobsen PB. Cognitive functioning after adjuvant chemotherapy and/or radiotherapy for early-stage breast carcinoma. Cancer. 2005;104(11):2499-507. | Review question related to radiotherapy impact with group comprising chemotherapy + radiotherapy |
| Du 2010 | Du XL, Xia R, Hardy D. Relationship between chemotherapy use and cognitive impairments in older women with breast cancer: findings from a large population-based cohort. Am J Clin Oncol. 2010;33(6):533-43. | Serious late stage complaints |
| Edwards 2018 | Edwards BJ, Zhang X, Sun M, Holmes HM, Ketonen L, Guha N, et al. Neurocognitive deficits in older patients with cancer. J Geriatr Oncol. 2018;9(5):482-7. | Outcomes measured prior to chemotherapy treatment |
| Freedman 2013 | Freedman RA, Pitcher B, Keating NL, Ballman KV, Mandelblatt J, Kornblith AB, et al. Cognitive function in older women with breast cancer treated with standard chemotherapy and capecitabine on Cancer and Leukemia Group B 49907. Breast Cancer Research and Treatment. 2013;139(2):607-16. | Outcome data not presented separately for chemotherapy treatment |
| Iconomou 2004 | Iconomou G, Mega V, Koutras A, Iconomou AV, Kalofonos HP. Prospective assessment of emotional distress, cognitive function, and quality of life in patients with cancer treated with chemotherapy. Cancer. 2004;101(2):404-11. | Outcomes not split based on cancer type |
| Janz 2007 | Janz NK, Mujahid M, Chung LK, Lantz PM, Hawley ST, Morrow M, et al. Symptom experience and quality of life of women following breast cancer treatment. J Womens Health (Larchmt). 2007;16(9):1348-61. | No measure of cognition reported |
| Lange 2019 | Lange, M.; Licaj, I.; Clarisse, B.; Humbert, X.; Grellard, J.M.; Tron, L.; Joly, F. Cognitive complaints in cancer survivors and expectations for support: Results from a web–based survey. Cancer Medicine 2019, 8, 2654-2663. | Outcome data not presented separately for chemotherapy treatment |
| Leinert 2019 | Leinert E, Schwentner L, Blettner M, Wöckel A, Felberbaum R, Flock F, et al. Association between cognitive impairment and guideline adherence for application of chemotherapy in older patients with breast cancer: Results from the prospective multicenter BRENDA II study. Breast J. 2019;25(3):386-92. | Outcome data not split based on treatment |
| Levkovich 2018 | Levkovich I, Cohen M, Alon S, Kuchuk I, Nissenbaum B, Evron E, et al. Symptom cluster of emotional distress, fatigue and cognitive difficulties among young and older breast cancer survivors: The mediating role of subjective stress. J Geriatr Oncol. 2018;9(5):469-75. | Outcomes not split based on cancer type |
| Li 2015 | Li J, Yu L, Long Z, Li Y, Cao F. Perceived cognitive impairment in Chinese patients with breast cancer and its relationship with post-traumatic stress disorder symptoms and fatigue. Psychooncology. 2015;24(6):676-82. | No prevalence data reported |
| Lycke 2017 | Lycke M, Lefebvre T, Pottel L, Pottel H, Ketelaars L, Stellamans K, et al. The distress thermometer predicts subjective, but not objective, cognitive complaints six months after treatment initiation in cancer patients. Journal of Psychosocial Oncology. 2017;35(6):741-57. | Outcomes not split based on cancer type |
| Lycke 2017 | Lycke M, Pottel L, Pottel H, Ketelaars L, Stellamans K, Van Eygen K, et al. Predictors of baseline cancer-related cognitive impairment in cancer patients scheduled for a curative treatment. Psycho-Oncology. 2017;26(5):632-9. | Outcomes not split based on cancer type |
| Lyon 2016 | Lyon DE, Cohen R, Chen H, Kelly DL, Starkweather A, Ahn HC, et al. The relationship of cognitive performance to concurrent symptoms, cancer- and cancer-treatment-related variables in women with early-stage breast cancer: a 2-year longitudinal study. J Cancer Res Clin Oncol. 2016;142(7):1461-74. | No prevalence data reported |
| Magnuson 2019 | Magnuson A, Lei L, Gilmore N, Kleckner AS, Lin FV, Ferguson R, et al. Longitudinal Relationship Between Frailty and Cognition in Patients 50 Years and Older with Breast Cancer. J Am Geriatr Soc. 2019;67(5):928-36. | No prevalence data reported |
| Mandelblatt 2018 | Mandelblatt JS, Small BJ, Luta G, Hurria A, Jim H, McDonald BC, et al. Cancer-related cognitive outcomes among older breast cancer survivors in the thinking and living with cancer study. Journal of Clinical Oncology. 2018;36(32):3211-22. | No prevalence data reported |
| Mandelblatt 2020 | Mandelblatt JS, Zhai W, Ahn J, Small BJ, Ahles TA, Carroll JE, et al. Symptom burden among older breast cancer survivors: The Thinking and Living With Cancer (TLC) study. Cancer. 2020;126(6):1183-92. | No prevalence data reported |
| Morin 2018 | Morin RT, Midlarsky E. Treatment With Chemotherapy and Cognitive Functioning in Older Adult Cancer Survivors. Arch Phys Med Rehabil. 2018;99(2):257-63. | Outcomes not split based on cancer type |
| Oh 2017 | Oh PJ. Predictors of cognitive decline in people with cancer undergoing chemotherapy. Eur J Oncol Nurs. 2017;27:53-9. | Outcomes not split based on cancer type |
| Pereira 2015 | Pereira S, Fontes F, Sonin T, Dias T, Fragoso M, Castro-Lopes JM, et al. Neurological complications of breast cancer: A prospective cohort study. Breast. 2015;24(5):582-7. | Outcome data not split based on treatment |
| Poppelreuter 2004 | Poppelreuter M, Weis J, Külz AK, Tucha O, Lange KW, Bartsch HH. Cognitive dysfunction and subjective complaints of cancer patients: A cross-sectional study in a cancer rehabilitation centre. European Journal of Cancer. 2004;40(1):43-9. | Outcomes not split based on cancer type |
| Pullens 2013 | Pullens MJ, De Vries J, Van Warmerdam LJ, Van De Wal MA, Roukema JA. Chemotherapy and cognitive complaints in women with breast cancer. Psychooncology. 2013;22(8):1783-9. | No prevalence data reported |
| Rick 2018 | Rick O, Reuß-Borst M, Dauelsberg T, Hass HG, König V, Caspari R, et al. NeuroCog FX study: A multicenter cohort study on cognitive dysfunction in patients with early breast cancer. Psycho-Oncology. 2018;27(8):2016-22. | Multifaceted intervention with all patients receiving radiation therapy |
| Schagen 2001 | Schagen SB, Hamburger HL, Muller MJ, Boogerd W, Van Dam FSAM. Neurophysiological evaluation of late effects of adjuvant high-dose chemotherapy on cognitive function. Journal of Neuro-Oncology. 2001;51(2):159-65. | Patients grouped based on chemotherapy dosage making comparison with other studies challenging |
| Scherwath 2006 | Scherwath A, Mehnert A, Schleimer B, Schirmer L, Fehlauer F, Kreienberg R, et al. Neuropsychological function in high-risk breast cancer survivors after stem-cell supported high-dose therapy versus standard-dose chemotherapy: Evaluation of long-term treatment effects. Annals of Oncology. 2006;17(3):415-23. | Multifaceted intervention with some patients receiving stem cell support |
| Shaffer 2012 | Shaffer VA, Merkle EC, Fagerlin A, Griggs JJ, Langa KM, Iwashyna TJ. Chemotherapy was not associated with cognitive decline in older adults with breast and colorectal cancer: findings from a prospective cohort study. Med Care. 2012;50(10):849-55. | No prevalence data reported |
| Vardy 2006 | Vardy J, Wong K, Yi QL, Park A, Maruff P, Wagner L, et al. Assessing cognitive function in cancer patients. Supportive Care in Cancer. 2006;14(11):1111-8. | Sample population selected based on CICI presence |
| Vardy 2008 | Vardy, J.L.; Xu, W.; Booth, C.M.; Park, A.; Dodd, A.; Rourke, S.; Dhillon, H.; Clarke, S.J.; Wagner, L.; Tannock, I.F. Relation between perceived cognitive function and neuropsychological performance in survivors of breast and colorectal cancer. Journal of Clinical Oncology 2008, 26, 9520-9520. | Sample population selected based on CICI presence |
| VonAh 2015 | Von Ah D, Tallman EF. Perceived cognitive function in breast cancer survivors: Evaluating relationships with objective cognitive performance and other symptoms using the functional assessment of cancer therapy - Cognitive function instrument. Journal of Pain and Symptom Management. 2015;49(4):697-706. | Sample population selected based on CICI concerns |
| Wazqar 2019 | Wazqar DY. Cognitive Dysfunction and Its Predictors in Adult Patients With Cancer Receiving Chemotherapy: A Cross-Sectional Correlational Study. J Nurs Res. 2019;27(6):e56. | Outcomes not split based on cancer type |
| Wefel 2004 | Wefel JS, Lenzi R, Theriault RL, Davis RN, Meyers CA. The cognitive sequelae of standard-dose adjuvant chemotherapy in women with breast carcinoma: results of a prospective, randomized, longitudinal trial. Cancer. 2004;100(11):2292-9. | Possible overlapping patient sample with later study included (Wefel 2010) |
| Wefel 2014 | Wefel JS, Kornet RL, Schagen SB. Systemically treated breast cancer patients and controls: an evaluation of the presence of noncredible performance. J Int Neuropsychol Soc. 2014;20(4):357-69. | Wrong aim since examining cognitive test reliability |
| Weis 2009 | Weis J, Poppelreuter M, Bartsch HH. Cognitive deficits as long-term side-effects of adjuvant therapy in breast cancer patients: 'Subjective' complaints and 'objective' neuropsychological test results. Psycho-Oncology. 2009;18(7):775-82. | Potentially biased sample population in rehabilitation centre |
| Zheng 2014 | Zheng Y, Luo J, Bao P, Cai H, Hong Z, Ding D, et al. Long-term cognitive function change among breast cancer survivors. Breast Cancer Res Treat. 2014;146(3):599-609. | No prevalence data reported |

**S5: Timeline of the cognitive assessments in relation to breast cancer treatment stages, illustrating grouping used for assimilation of data across the studies (n represents patient numbers)**

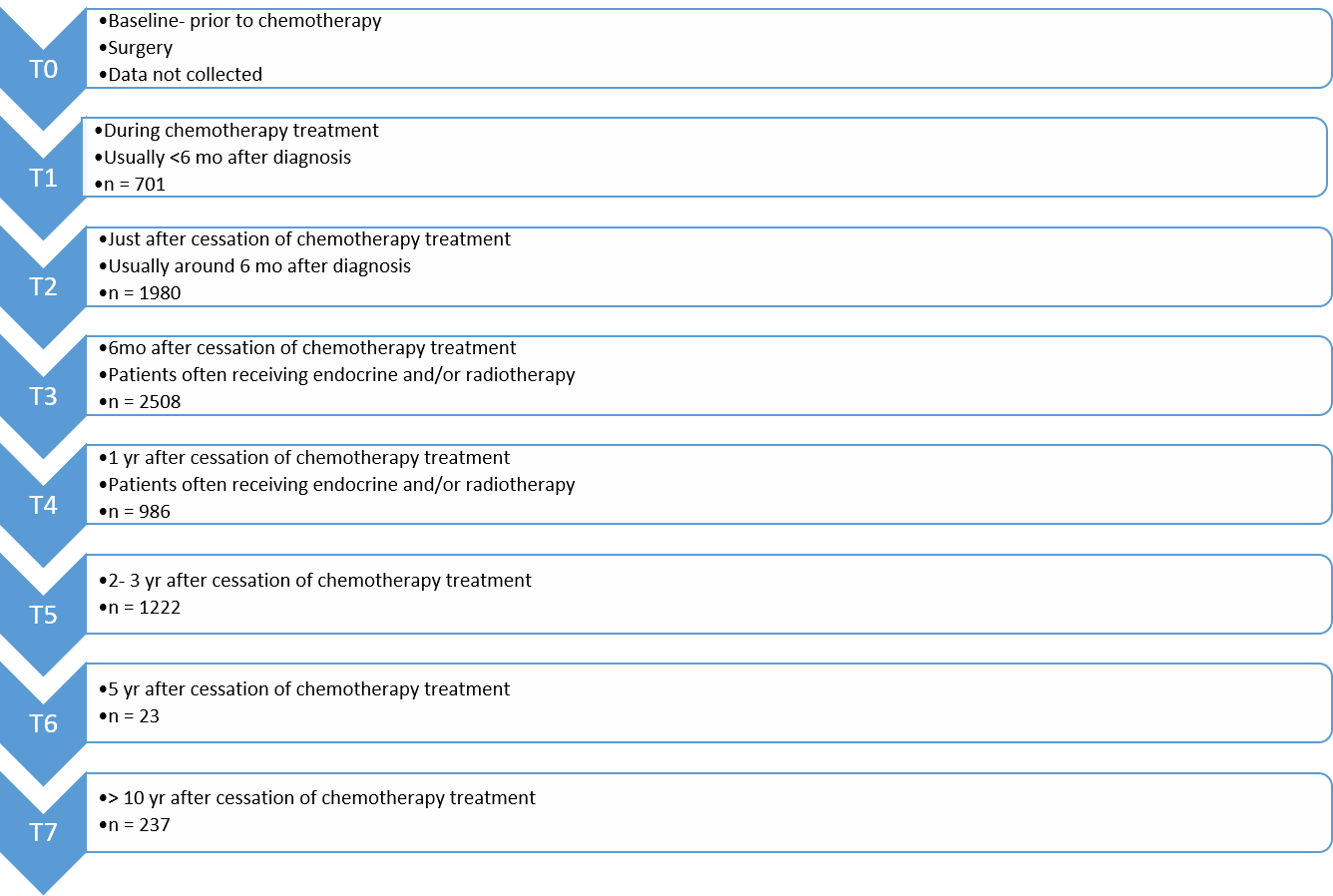

**S6: Details of the methods employed to assess cognitive impairment in the included studies. Adapted from Collins et al 2009. [84]**

| Cognitive Domains and Tests | Description |
| --- | --- |
| Self-reported Cognitive Function Tests | |
| Broadbent (Broadbent) cognitive failures questionnaire [54] | A series of 25 questions relating to lapses in attention in everyday  life, such as forgetting what the person went into a room to do.  Questions are rated on a five-point scale ranging from 0-‘never’ to  5-‘very often. |
| Functional Assessment of Cancer Therapy - Cognitive Function (FACT-Cog) [55] | Likert scale response test which considers patients perceived cognitive impairments, impact of the perceived impairments on quality of life, comments from others and perceived cognitive abilities. |
| Patient’s Assessment of Own Functioning Inventory (PAOFI)[56] | Validated test with 33 questions based on four cognitive domains: memory, higher-level cognition (executive function), language and communication, and motor-sensory perception. Each item is rated on a 6-point Likert scale. |
| Quality of Life in Adult Cancer Survivors (QLACS) [57] | This tool covers five cancer-specific areas, as well as seven additional areas that are cancer relevant but not limited to cancer. It includes questions relating to concentration, forgetting, attention and remembering things. |
| Squire (Squire) Memory Self-Rating Questionnaire [58] | 18-item questionnaire covering new learning, working memory and remote memory. Patients report on a numeric scale based on perceived difference in criteria from previously. |
| Short Cognitive Screening | |
| Doors and People Test | A test of memory developed as a battery of tests with duration of around 45 minutes. [59] It consists of four categories: doors, people, shapes and names which test visual recognition, verbal recall, visual recall and verbal recognition respectively. The test has good face validity, but may be culturally biased and has complex scoring. |
| Headminder computerized test [60] | A 30-minute, internet-based, computerized test that automates collection of objective measures of automated, objective measures of attention, memory, response speed, and processing speed. Often used for an initial evaluation (triage) of cognitive functioning. |
| Mini-Mental State Examination (MMSE) [61] | Short test of duration between 5 and 10 minutes which evaluates registration, attention and calculation, recall, language, orientation to place, and ability to follow simple commands. |
| Montreal Cognitive Assessment (MoCA) [62] | A 30-point test, which can be administered in approximately 10 min. The test covers domains of attention and working memory, short-term memory recall, visuospatial abilities, language abilities, and executive function. Suggested to be more sensitive than the MMSE at detecting mild cognitive impairment. |
| The High Sensitivity Cognitive  Screen (HSCS) [63] | 20-minute interview-based test. Assesses a range of functions across various cognitive domains, including memory, language, attention and concentration, visual and motor skills, spatial perception, self-regulation and executive functioning. |
| Objective Neuropsychological Tests | |
| *Cognitive Domains* | |
| Executive Function | An umbrella term describing various cognitive processes which may include problem solving, planning, organisational skills, and aspects of attention. Typical tests used include the Stroop Color-Word Test [64], Trail Making Test B [65], Wisconsin Card Sorting Test, [66] Word Fluency Test [67], and Paced Auditory Serial Addition Test (PASAT).[68] |
| Language Function | Tests impairment to language, which may include speech, reading and writing. Commons tests include the Boston Naming Test [69] and Controlled Oral Word Association Test. [70] |
| Motor | Motor function tests assess the ability to precisely move muscles to perform a specific act.  Deficits in fine motor function may occur in CICI which may be reflected in time to perform a manual task. Common tests include Fepsy Finger Tapping [71] and the Grooved Pegboard [72]. |
| Processing Speed | Processing speed is assumed to relate to ability to learn new information. This task requires receiving the information, understanding it and generating a reaction. Many of these tests assess reaction times to various stimuli. Common tests include the Attentive matrices test [73], Conner ’s Continuous Performance Test [74], Digit Symbol Coding, WAIS-III [75]  and Trail Making Test A [65]. |
| Verbal Learning and Memory | These tests are usually highly sensitive to cognitive decline. Tests usually measure immediate memory span, new verbal learning, susceptibility to modification, memory recognition and retention of information after a delay. Common tests include the California Verbal Learning Test [76], Hopkins Verbal Learning Test [77], and Logical Memory II [78]. |
| Visual Learning and Memory | Measures visual (non-verbal) ability to learn and remember as one way of assessing memory performance. They measure a person’s ability to hold visual images in their mind. Tests may present figures, images of faces, or pictures. Common tests include the Brief Visuospatial Memory Test Revised [79], Family Pictures II [78], and the Rey Visual Learning Test (RVLT) [80] |
| Visuo-Spatial Function | Visuospatial function involves the identification of a stimulus and its location and tests cover visual perception, construction and integration. Commons tests include the Block Design [81] and the Design Organization Test[82]. |
| Working Memory | Working memory represents the ability to hold information temporarily. It is important for reasoning and to guide decision-making. Common tests employed are the Arithmetic [81], Consonant Trigrams [83], Digit Span [81] and Letter-Number Sequencing [81]. |

##### S7:Risk of bias summary: review authors' judgements about each risk of bias item for each included study

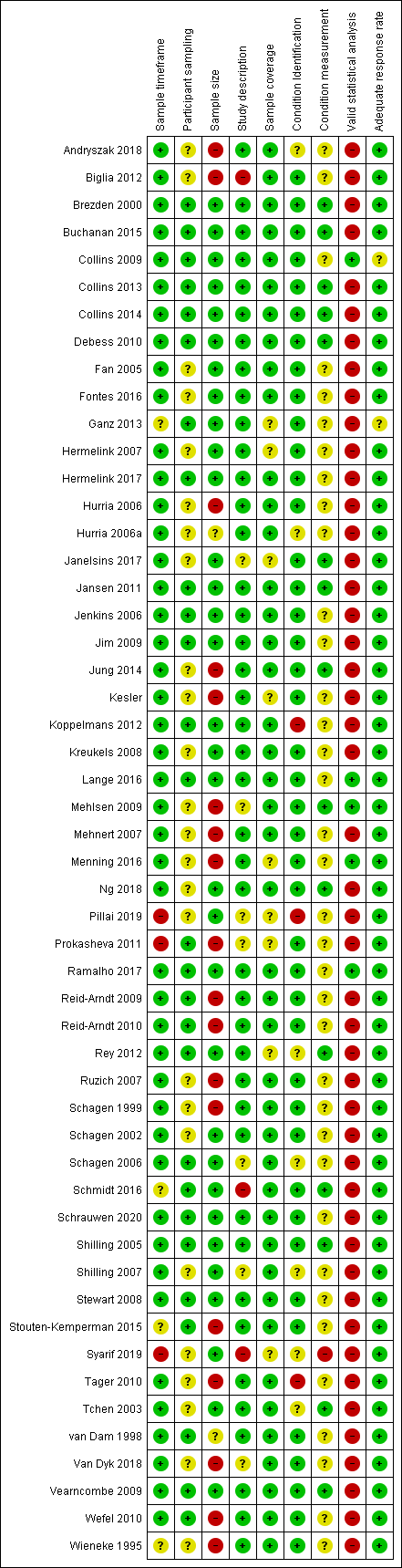

### **S8: Sensitivity Analyses**

| **Sensitivity analysis performed** |  |  |  |
| --- | --- | --- | --- |
|  | T1 **(*I*^2^=73%)** | T2 (**(*I*^2^=90%)** | T4 (***I*^2^=89%)** |
| Removal of each study in turn | Removal of Jansen 2011 reduced *I*^2^ by 10% to 63% (prevalence 31%) | Minimal effect | Minimal effect |
| No transformation | Reduced I^2^ to 70% | Minimal effect | Minimal effect |
| Quality effects model applied | Minimal effect | Minimal effect | Minimal effect |
| Fixed effects model | Minimal effect | Heterogeneity unchanged, pooled prevalence drop of 4%, | Heterogeneity unchanged, pooled prevalence drop of 5%, |

**Table of sensitivity analyses performed when *I*^2^ ≥ 70% and their impacts on results**

### **S9: Doi plots.** The LFK index is reported as a value with the interpretation that an index within ±1, from ±1 to ±2, and > ± 2 represent no asymmetry, minor asymmetry and major asymmetry, respectively. [39]

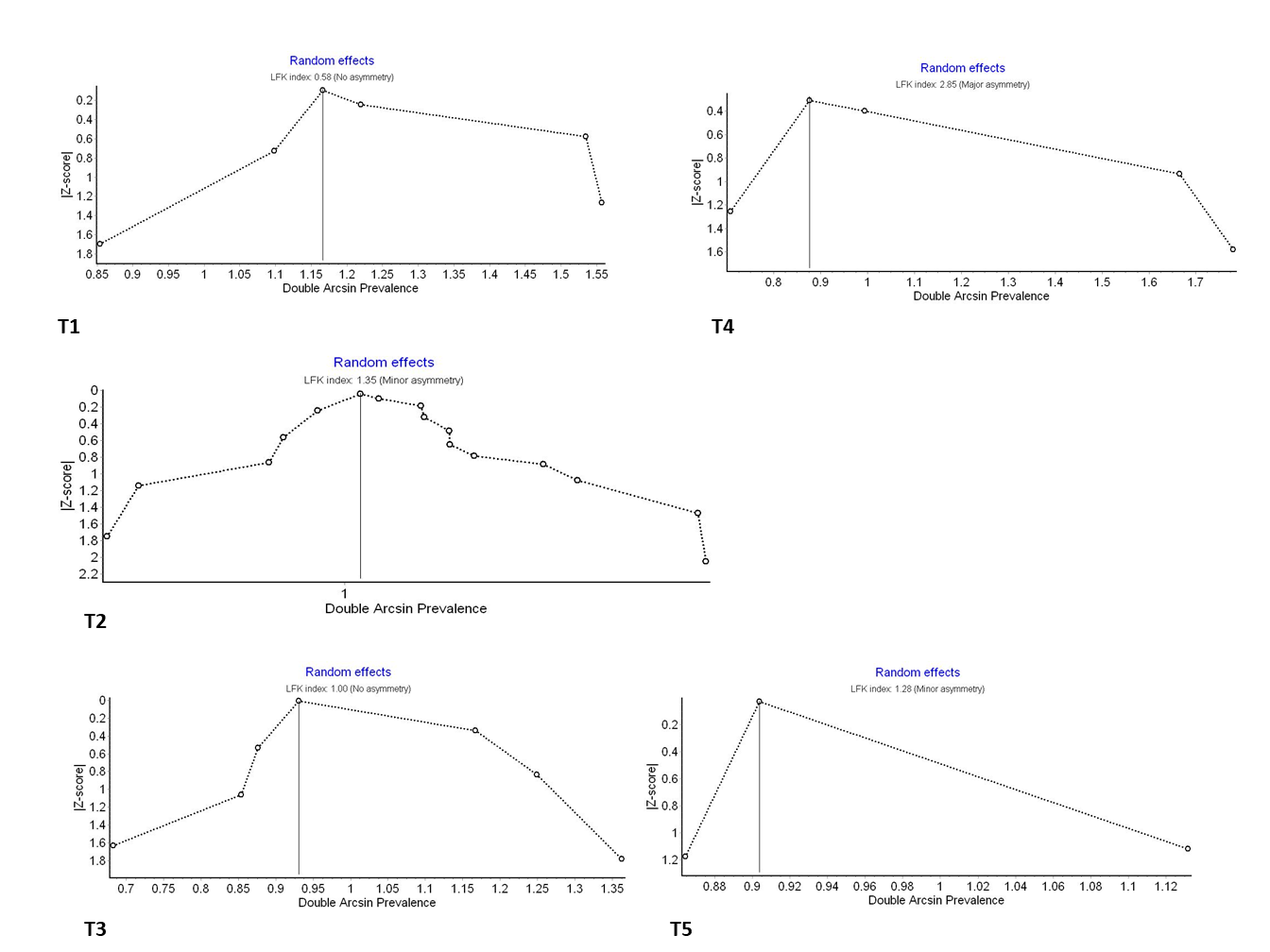
